# Genetic dissection of *HLA-DRB1*15:01* and XL9 region variants in Japanese patients with systemic lupus erythematosus: Primary role for *HLA-DRB1*15:01*

**DOI:** 10.1101/2023.04.05.23288103

**Authors:** Aya Kawasaki, Premita Ari Kusumawati, Yuka Kawamura, Yuya Kondo, Makio Kusaoi, Hirofumi Amano, Yasuyoshi Kusanagi, Kenji Itoh, Takashi Fujimoto, Naoto Tamura, Hiroshi Hashimoto, Isao Matsumoto, Takayuki Sumida, Naoyuki Tsuchiya

## Abstract

**Objective:** Major histocompatibility complex strongly contributes to susceptibility to systemic lupus erythematosus (SLE). In the European populations, *HLA-DRB1*03:01* and *DRB1*15:01* are susceptibility alleles, but *C4* locus was reported to account for the association of *DRB1*03:01*. With respect to *DRB1*15:01*, strong linkage disequilibrium (LD) with a variant rs2105898T in the XL9 region, located between *DRB1* and *DQA1* and regulates HLA-class II expression levels, was reported; however, the causative allele remains to be determined. Leveraging the genetic background of the Japanese population, where *DRB1*15:01* and *DRB1*15:02* are commonly present and only *DRB1*15:01* is associated with SLE, this study aimed to distinguish the genetic contribution of *DRB1*15:01* and XL9 variants.

**Methods:** Among the XL9 variants, two (rs2105898 and rs9271593) previously associated variants in the European populations and two (rs9271375 and rs9271378) which showed a trend towards association in a Japanese genome-wide association study were selected. Associations of the XL9 variants and *HLA-DRB1* were examined in 442 Japanese SLE patients and 779 controls. Genotyping of the XL9 variants were performed by TaqMan SNP Genotyping Assay and direct sequencing. *HLA-DRB1* alleles were determined by polymerase chain reaction-reverse sequence-specific oligonucleotide probes.

**Results:** Among the XL9 variants, associations of rs2105898T and rs9271593C were replicated in the Japanese population. However, these associations became no longer significant when conditioned on *DRB1*15:01*. In contrast, the association of *DRB1*15:01* remained significant after conditioning on the XL9 variants.

**Conclusion:** In the Japanese population, *HLA-DRB1*15:01* was found to be primarily associated with SLE, and to account for the apparent association of XL9 region.

**WHAT IS ALREADY KNOWN ON THIS TOPIC:**

- The association of *HLA-DRB1*03:01* with susceptibility to systemic lupus erythematosus (SLE) was reported to be secondarily caused by linkage disequilibrium (LD) with copy number reduction of *C4*, which has the primary role.
- A possibility has been hypothesized that the association of *HLA-DRB1*15:01* with SLE may possibly be caused by LD with XL9 region variants, associated with expression levels of HLA-class II; however, due to strong LD between *DRB1*15:01* and XL9 variants, this hypothesis could not be addressed in the European populations.

**WHAT THIS STUDY ADDS:**

- In the Japanese population, two common *DRB1*15* alleles, *DRB1*15:01* and *DRB1*15:02*, are present, both in LD with XL9 variants. However, only *DRB1*15:01* is associated with SLE.
- Leveraging the population difference in the genetic background, we demonstrated that *DRB1*15:01*, rather than XL9 region variants, is primarily associated with SLE in the Japanese population.

**HOW THIS STUDY MIGHT AFFECT RESEARCH, PRACTICE OR POLICY:**

- This study provides us with critical information in understanding the respective roles of *HLA* genes and their regulatory regions in the development of SLE.
- This study also shows the usefulness of association studies in multiple populations with different genetic backgrounds in the identification of causative variants.

## INTRODUCTION

Systemic lupus erythematosus (SLE) is a systemic autoimmune disease caused by a combination of multiple genetic and environmental factors. Genetic studies including genome-wide association studies (GWAS) have identified approximately 180 susceptibility loci for SLE.[1-3] In the European and the African populations, the most significant association was detected in the major histocompatibility complex (*MHC*) region,[2] where complement *C4* and human leukocyte antigen (*HLA*) loci are located. In the Asian populations, the *GTF2I-NCF1* region was most strongly associated with SLE,[3-5], where *NCF1* p.Arg90His (rs201802880) has been reported to be the causative variant.[5-7] Nevertheless, *MHC* remains one of the top susceptibility regions also in the Asian populations.[3,4]

Among the *HLA* alleles, *HLA-DRB1*03:01* and *DRB1*15* have been established as susceptibility alleles to SLE.[8,9] Although *DRB1*03:01* is associated with SLE in the European populations, *DRB1*03:01* is rare in the East Asian populations, and significant association of *DRB1*03:01* is not always detected.[10,11] In contrast, the association of *DRB1*15:01* is shared between the European and the East Asian populations.[8-11] Further, *DRB1*15:03* and *DRB1*15:02* were associated with SLE in the African and the Southeast Asian populations, respectively.[12-14]

Identification of causative allele(s) in the *MHC* region has been challenging, mainly because of the presence of numerous potentially functional variants of *HLA* and non-*HLA* genes, and linkage disequilibrium (LD) that extends from the *MHC class I* to *class II* regions. Thus, each *DRB1* allele is a part of a haplotype that carries multiple potentially functional variants.[8] Most importantly, *C4* genes, *C4A* and *C4B*, reside in the *MHC class III* region, and have a copy number variation. A lower copy number of *C4*, especially *C4A*, has been shown to confer risk to SLE.[15,16] In the European populations, due to the strong LD between *C4A* deficient allele and *DRB1*03:01*, it was impossible to distinguish the genetic contribution of these two loci in the development of SLE. However, leveraging the genetic background of the African-American population, where the LD between *C4A* deficiency and *DRB1*03:01* is low, Kamitaki et al. successfully demonstrated that *C4A* deficiency has the primary role, while the association of *DRB1*03:01* was attributable to the LD with *C4*.[15]

With respect to the other risk haplotype carrying *DRB1*15:01*, they also detected an association of a single nucleotide variant (SNV), rs2105898T, in the XL9 region between *HLA-DRB1* and *DQA1* loci, both in the European and the African American populations, which was independent of *C4*.[15] The XL9 region risk variants were associated with expression levels of HLA class II molecules. Raj et al. also demonstrated the association of a SNV in the XL9 region, rs9271593,[17] which is in LD with rs2105898. The XL9 risk variants were in strong LD with *DRB1*15:01* in the European population; thus, the causative allele among them has not been inferred [15,17].

In the European and the African populations, *HLA-DRB1*15:01* and *DRB1*15:03* accounts for the majority of *HLA-DRB1*15*, respectively. In contrast, both *DRB1*15:01* and *DRB1*15:02* are commonly present in the Japanese population, of which only *DRB1*15:01* is associated with susceptibility to SLE.[11] By leveraging genetic background of the Japanese population, this study was carried out to test whether the association signal of *HLA-DRB1*15:01* and the XL9 region variant can be distinguished in the Japanese population.

## SUBJECTS AND METHODS

### Patients and controls

A total of 1,221 Japanese individuals were analyzed, including 442 patients with SLE and 779 healthy controls. Characteristics of the patients and controls are summarized in Table 1. Patients’ genomic DNA samples were recruited at University of Tsukuba, the University of Tokyo, Juntendo University, Nara Medical University, and National Defense Medical College. The patients were diagnosed with SLE according to the 1997 American College of Rheumatology revised criteria for the classification of SLE.[18] In addition, 515 genomic DNA samples were obtained from healthy volunteers at University of Tsukuba, the University of Tokyo, and Juntendo University. Aside from that, 264 genomic DNAs were purchased from the National Institute of Biomedical Innovation (Osaka, Japan).

**Table 1.** Characteristics of the patients with SLE and healthy controls

|  | SLE (n=442) | Healthy controls (n=779) |
| --- | --- | --- |
| Male/female | 35/399* | 304/475 |
| Mean age ± SD (years) | 42.0 ± 14.0 | 36.3 ± 11.5 |
| Age of onset (n=377) |  |  |
| < 20 years, n (%) | 81 (21.5) | NA |
| Renal disorder (n=376) |  |  |
| Present, n (%) | 203 (54.0) | NA |
| Anti-dsDNA antibody (n=363) |  |  |
| Present, n (%) | 298 (82.1) | NA |
| Anti-Sm antibody (n=362) |  |  |
| Present, n (%) | 75 (20.7) | NA |

SLE, systemic lupus erythematosus; SD, standard deviation; NA, not applicable.
\*data was not available for eight patients with SLE.

### Genotyping

Genotyping of rs2105898 and rs9271593 was performed by Custom TaqMan SNP Genotyping Assays (Thermo Fisher Scientific, Waltham, MA, USA) using the QuantStudio 5 Real-Time PCR System (Thermo Fisher Scientific). Genotypes of rs9271375 and rs9271378 were determined by Sanger sequencing. A genomic region surrounding rs9271375 and rs9271378 was amplified and cycle sequencing reaction was conducted using a SupreDye v3.1 Cycle Sequencing Kit (Edge BioSystems, San Jose, CA, USA). Subsequently, sequencing was performed using 3500 Genetic Analyzer (Thermo Fisher Scientific). Primers and probes used in these assays were listed in Supplementary Table S1(Online Supplementary Materials). *HLA-DRB1* alleles were determined at four-digit resolution by polymerase chain reaction-reverse sequence-specific oligonucleotide probes using a WAKFlow HLA typing kit (Wakunaga Pharmaceutical Co., Ltd., Osaka, Japan). The genotyping results of each subject are available in Supplementary Data (Online Supplementary Materials).

### GWAS data on SLE in a Japanese population

GWAS data on 317 Japanese patients with SLE and 175,937 Japanese controls previously reported by Sakaue et al.[19] was retrieved from the NBDC Human Database (https://humandbs.biosciencedbc.jp/en/, Dataset ID: hum0197.v3.gwas.v1, accessed 29 Nov 2022). Data on variants in a region between *HLA-DRB1* and *HLA-DQA1* encompassing XL9 was extracted.

### Statistical methods

Statistical analyses were carried out by logistic regression analysis with an additive model using the R software version 3.5.2. Association was tested for SLE and its subsets. Correction for multiple testing (four SNVs) was performed by the false discovery rate (FDR) method based on the Benjamini-Hochberg procedure. Significance level was set at FDR Q<0.05. The association of haplotypes formed by *HLA-DRB1* and the XL9 variants with SLE was tested by permutation test (the number of permutations was 10,000,000) using Haploview 4.2 software (Broad Institute, Cambridge, MA). In LD analysis, *r*^*2*^ values were calculated by the Haploview software. Power calculation was done using Quanto ver 1.2.4. On the basis of the sample size (442 cases and 779 controls) and previously reported effect size in each study,[15,17,19] this study is assumed to have the power of 73% (rs2105898), 99% (rs9271593), 82% (rs9271375) and 68% (rs9271378) to detect the association with Bonferroni corrected statistical significance, α =0.0125.

## RESULTS

### Association of XL9 variants with SLE

Four XL9 variants were selected for the association analysis between the XL9 region and SLE in the Japanese population. Two of them, rs2105898 and rs9271593, were previously reported to be associated with SLE in the European populations,[15,17] and were chosen to examine whether they are also associated in the Japanese population. In addition, in view of differences in the genomic configuration of MHC region between the European and the Asian populations, we employed the GWAS data on Japanese SLE reported by Sakaue et al.[19] available at the NBDC Human Database (NBDC Dataset ID: hum0197.v3.gwas.v1 https://humandbs.biosciencedbc.jp/en/). In this GWAS, although no XL9 variants reached genome-wide significance, two SNVs, rs9271375 and rs9271378, showed a trend towards association with SLE (Supplementary Figure S1 and Supplementary Table S2, Online Supplementary Materials). Thus, the association of rs9271375 and rs9271378 was also tested in this study.

When LD status among the four XL9 variants were tested, LD was observed between rs2105898 and rs9271593 (r^2^=0.676) (Supplementary Figure S2, Online Supplementary Materials). As shown in Table 2, rs2105898T and rs9271593C were significantly associated with risk of SLE after correction for multiple comparisons (FDR Q=0.0067 and 0.037, respectively). Thus, these SNVs, previously associated in the European populations,[15,17] were found to be associated also in the Japanese. With respect to rs9271375 and rs9271378, the same trend towards the association as observed in the previous Japanese GWAS[19] was also detected in our subjects. However, the difference did not reach statistical significance in our sample set (FDR Q=0.051 for both SNVs) (Table 2).

**Table 2.** Association of XL9 variants with SLE

|  | genotype |  |  | effect allele | additive model |  |  |
| --- | --- | --- | --- | --- | --- | --- | --- |
|  |  |  |  |  | P | FDR Q* | OR (95% CI) |
| rs9271375A/G | A/A | A/G | G/G | A |  |  |  |
| SLE (n=442) | 39 (8.8) | 172 (38.9) | 231 (52.3) | 250 (28.3) | 0.051 | 0.051 | 0.84 (0.70-1.00) |
| Controls (n=779) | 78 (10.0) | 344 (44.2) | 357 (45.8) | 500 (32.1) |  |  | referent |
| rs9271378G/A | G/G | G/A | A/A | G |  |  |  |
| SLE (n=442) | 7 (1.6) | 84 (19.0) | 351 (79.4) | 98 (11.1) | 0.038 | 0.051 | 0.77 (0.59-0.98) |
| Controls (n=779) | 16 (2.1) | 187 (24.0) | 576 (73.9) | 219 (14.1) |  |  | referent |
| rs2105898T/G | T/T | T/G | G/G | T |  |  |  |
| SLE (n=442) | 48 (10.9) | 174 (39.4) | 220 (49.8) | 270 (30.5) | 0.0017 | 0.0067 | 1.34 (1.12-1.61) |
| Controls (n=779) | 46 (5.9) | 291 (37.4) | 442 (56.7) | 383 (24.6) |  |  | referent |
| rs9271593C/T | C/C | C/T | T/T | C |  |  |  |
| SLE (n=442) | 63 (14.3) | 202 (45.7) | 177 (40.0) | 328 (37.1) | 0.018 | 0.037 | 1.23 (1.04-1.46) |
| Controls (n=779) | 83 (10.7) | 338 (43.4) | 358 (46.0) | 504 (32.3) |  |  | referent |

SLE, systemic lupus erythematosus; FDR; false discovery rate, OR, odds ratio; CI, confidence interval.
\*Significance level was set at FDR Q<0.05.

We next tested whether the XL9 variants were associated with the age of onset and presence or absence of renal disorder, anti-dsDNA antibodies and anti-Sm antibodies by a case-case study. As shown in Supplementary Table S3 (Online Supplementary Materials), no significant association was detected with the SLE subsets.

### Linkage disequilibrium between HLA-DRB1 and XL9 variants

*HLA-DRB1*15* is an established susceptibility allele group to SLE. *DRB1*15:01* and *DRB1*15:03* are predominant in the European and the African populations, respectively, while both *DRB1*15:01* and *DRB1*15:02* alleles are common in the East Asian populations. The XL9 region is located between the *HLA-DRB1* and *DQA1* loci, and LD is observed between *DRB1*15* and XL9 variants. We retrieved genotype data on *HLA-DRB1*, rs2105898 and rs9271593 in the Utah residents (CEPH) with Northern and Western European ancestry (CEU), Japanese in Tokyo, Japan (JPT), Han Chinese in Beijing (CHB), and Yoruba in Ibadan, Nigeria (YRI) from the International Genome Sample Resource (2018 data, https://www.internationalgenome.org/category/hla/, accessed 28 Jun 2022) and the Ensembl database (Release 106, https://asia.ensembl.org/index.html, accessed 28 Aug 2022).[20] and calculated *r*^*2*^ values between each *DRB1*15* allele and XL9 variant (Supplementary Table S4, Online Supplementary Materials). *DRB1*15:01* and *DRB1*15:03* are associated with SLE in the European- and the African-ancestry populations, respectively, [8,9,12,15] and rs2105898T is associated with SLE in both populations;[15] however, due to strong LD, association signals from *DRB1*15* and XL9 variants have not been clearly discriminated in these populations.

In contrast, in the Japanese population, moderate LD was observed between *DRB1*15* (*\*15:01* and *\*15:02*) and XL9 variants (rs2105898 and rs9271593) (Table 3, Supplementary Tables S4, S5 and Figure S2, Online Supplementary Materials). Although both *DRB1*15:01* and *DRB1*15:02* are commonly present in the Japanese population and in LD with the XL9 variants, *DRB1*15:01*, but not *DRB1*15:02*, was associated with SLE (Table 3), as previously reported.[11] Therefore, we thought that genetic dissection of *DRB1*15:01* and XL9 variants might be possible by analyzing the Japanese subjects.

**Table 3.**
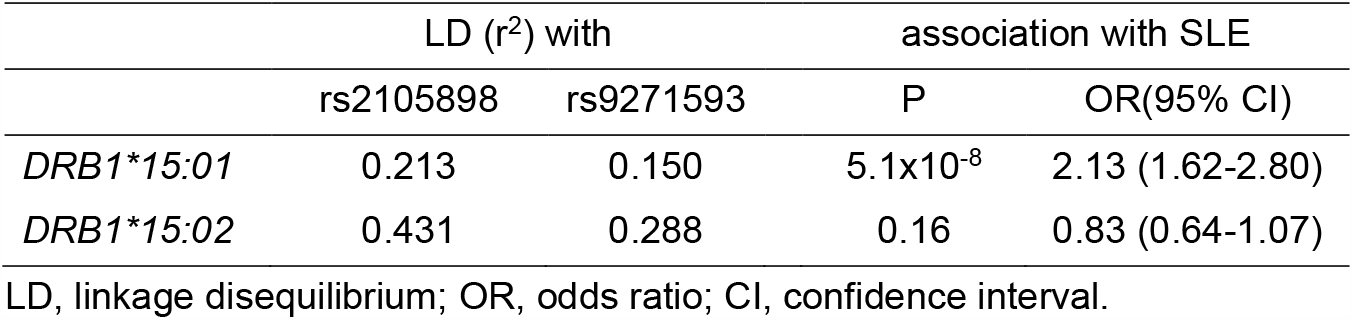
Linkage disequilibrium between *DRB1*15* alleles and XL9 variants and association between *DRB1*15* alleles and susceptibility to SLE.

### The primary role for HLA-DRB1*15:01

To address this issue, we conducted a conditional logistic regression analysis to examine whether *DRB1*15:01* and the XL9 variants are independently associated with SLE. Table 4 shows that the association of *DRB1*15:01* remained significant after conditioning on rs2105898 (P_conditional_=7.6×10^−6^) or rs9271593 (P_conditional_=8.7×10^−7^), while that of rs2105898 and rs9271593 became no longer significant when conditioned on *DRB1*15:01* (rs2105898, P_conditional_= 0.83, rs9271593, P_conditional_= 0.93).

**Table 4.** Conditional logistic regression analysis of *HLA-DRB1*15:01* and XL9 variants

|  | unconditional |  | conditional on *DRB1\*15:01* |  | conditional on rs2105898 |  | conditional on rs9271593 |  |
| --- | --- | --- | --- | --- | --- | --- | --- | --- |
|  | P | OR<br>(95% CI) | P | OR<br>(95% CI) | P | OR<br>(95% CI) | P | OR<br>(95% CI) |
| *HLA-DRB1\*15:01* | 5.1x10<sup>-8</sup> | 2.13<br>(1.62-2.80) | NA | NA | 7.6x10<sup>-6</sup> | 2.09<br>(1.52-2.89) | 8.7x10<sup>-7</sup> | 2.14<br>(1.58-2.90) |
| rs2105898T | 0.0017 | 1.34<br>(1.12-1.61) | 0.83 | 1.02<br>(0.82-1.28) | NA | NA | 0.035 | 1.44<br>(1.03-2.05) |
| rs9271593C | 0.018 | 1.23<br>(1.04-1.46) | 0.93 | 0.99<br>(0.81-1.20) | 0.61 | 0.92<br>(0.66-1.27) | NA | NA |

OR, odds ratio; CI, confidence interval; NA, not applicable.

Haplotypes formed by *HLA-DRB1*15:01* and the XL9 variants were also tested for their association with SLE (Table 5). *DRB1*15:01*-rs2105898T-rs9271593C haplotype was significantly increased in SLE compared with healthy controls (permutation P = 1.0 × 10^−7^); however, the haplotype with rs2105898T and rs9271593C risk alleles, but without *DRB1*15:01*, was not associated with SLE. Taken together, these results suggested that the association of *DRB1*15:01* has the primary role in the susceptibility to SLE, and the association of the XL9 variants is secondarily caused by LD with *DRB1*15:01*.

**Table 5.** Association between SLE and haplotypes formed by *HLA-DRB1* and XL9 variants, rs2105898 and rs9271593

| haplotype |  |  | Frequency (%) |  | Permutation test* |
| --- | --- | --- | --- | --- | --- |
| *HLA-DRB1* | rs2105898 | rs9271593 | SLE | controls | P |
| *15:01 | T | C | 13.5 | 6.6 | 1.0x10<sup>-7</sup> |
| others | T | C | 17.0 | 18.0 | 0.95 |
| others | G | C | 6.6 | 7.8 | 0.65 |
| others | G | T | 62.6 | 67.5 | 0.041 |

\* The number of permutations: 10,000,000

## DISCUSSION

Identification of the causative variant(s) among the *MHC* region remains challenging, due to the presence of multiple potentially functional variants and extensive LD. With respect to *DRB1*03:01* haplotype in SLE, Kamitaki et al. reported a primary role for reduced copy number of *C4A* based on the results from the African-American population.[15] However, their study did not determine the causative allele on the *DRB1*15* haplotype, because of the tight LD with the XL9 region SNV which might regulate expression levels of *HLA class II* alleles, in the European population. In this study, leveraging the genetic background of the Japanese population, we were able to present evidence for the primary role of the *DRB1*15:01* allele, rather than the XL9 region SNVs, for the susceptibility to SLE.

The XL9 region shows high histone acetylation levels and interacts with the promoters of *HLA-class II* genes, including *HLA-DRB1* and *DQA1*. Thus the XL9 region was predicted to impact on transcriptional regulation of nearby *HLA* genes.[21,22] In fact, the risk alleles for SLE, rs2105898T and rs9271593C, were shown to be associated with expression of *HLA-class II* genes.[15,17] Whole blood eQTL data from the Genotype-Tissue Expression (GTEx) Portal V8 (https://gtexportal.org/home/, accessed 13 Mar 2023) are shown in Supplementary Table S6 (Online Supplementary Materials). Kamitaki et al. also reported that a binding site for a transcription factor ZNF143 was disrupted by substituting rs2105898G with T.[15] These data support the functional significance of XL9 variants.

Nevertheless, concerning the susceptibility to SLE, our conditional logistic regression analysis showed that the association of XL9 variants was no longer significant when conditioned on *DRB1*15:01*, whereas the association of *DRB1*15:01* remained significant after conditioning on XL9 variants, suggesting that *DRB1*15:01*, rather than XL9 variants, may be the causative susceptibility allele, at least in the Japanese population.

Consistent with this, although XL9 variants were in LD with both *DRB1*15:01* and *DRB1*15:02*, only *DRB1*15:01* was significantly associated with SLE susceptibility (Table 3). There is only a single amino acid difference at position 86 between *DRB1*15:01* (86Val) and *DRB1*15:02* (86Gly). This amino acid sequence difference affects the peptide binding motif (MHC Motif Viewer (https://services.healthtech.dtu.dk/services/MHCMotifViewer/Home.html)[23] and T cell receptor repertoire selection.[24] Furthermore, *DRB1*15:01* was recently reported to be associated with hypomethylation and increased expression of *HLA-DRB1* in monocytes.[25] Taken together, it appears more plausible that *DRB1*15:01* plays a primary role in the susceptibility to SLE through its effect on antigenic peptide specificity and/or T cell repertoire selection. On the other hand, the possible role for XL9 variants might not be entirely excluded. As shown in Table 5, a haplotype carrying *DRB1*15:01* contains rs2105898T and rs9271593C, and thus it might be possible that by having regulatory XL9 variants together with *DRB1*15:01* on the same chromosome, the effect of *DRB1*15:01* could be enhanced as compared with having the non-risk variants of XL9.

Recently, Wang et al. examined the genetic correlation for SLE between the European and the Chinese populations.[26] The transancestral genetic-effect correlation (r_ge_) was increased from 0.64 to 0.78 when the variants in the *HLA* region were removed from the analysis, suggesting a genetic difference in the *HLA* region between the European and the Chinese populations. The possibility that the role of the XL9 region may also be different among European, African and Asian populations cannot be excluded.

In view of population-specific differences in the genomic configuration of the MHC region, the possibility that XL9 variants other than rs2105898T and rs9271593C play a causal role cannot be excluded. Therefore, to comprehensively investigate an association of XL9 variants with SLE, we accessed the previous GWAS data on Japanese SLE,[19] and included two SNVs with a tendency towards association, rs9271375 and rs9271378, into our analysis. In our case-control set, although a trend towards association of rs9271378 was observed, the statistical significance did not stand correction for multiple testing. These SNVs were not in LD with *DRB1* alleles associated with SLE in the Japanese population, including *HLA*-*DRB1*15* alleles (Supplementary Table S5 and Figure S2, Online Supplementary Materials); therefore, the possibility that future large-scale studies might lead to identification of XL9 variant(s) with an independent effect from *DRB1*15:01* cannot be excluded.

Limitations of this study include a relatively small sample size. Although our sample size is assumed to have 68% or larger power to detect statistical significance based on previously reported effect size for each SNV,[15,17,19] studies in larger sample sizes may be necessary to comprehensively evaluate the independent role of XL9 region. In addition, this study does not provide any explanation on the significant association of *DRB1*15:02* in Southeast Asian populations.[13,14] Future fine mapping studies on these populations are required.

In summary, we replicated the association of the XL9 variants with SLE in a Japanese population and confirmed the observation reported in the European and the African populations. Taking advantage of the genetic background of the Japanese population, the association of the XL9 variants was suggested to be attributable to LD with *HLA-DRB1*15:01*, which may play a causative role. These observations further emphasize the transethnic studies’ importance in finely mapping causative variants.

## Data Availability

All data relevant to the study are included in the article or uploaded as Online Supplementary Materials.

## Ethics

This study was reviewed and approved by the of University of Tsukuba Institute of Medicine Ethics Committee (approval ID: 122(1), 123, 268). In addition, this study was also approved by the Ethics Committees of the following institutes. which participated in the collaboration and/or recruitment of the subjects: the University of Tokyo, Nara Medical University, Juntendo University and National Defense Medical College.

This study was conducted in accordance with the principles of the Declaration of Helsinki and Ethical Guidelines for Human Genome/Gene Analysis Research implemented by the Ministry of Education, Culture, Sports, Science and Technology, Ministry of Health, Labour and Welfare, and Ministry of Economy, Trade and Industry, of Japan. Written informed consent was obtained from each participant.

## Contributors

Dr. Kawasaki, Dr. Kusumawati and Dr. Tsuchiya designed the study, interpreted the data, and wrote the manuscript. Dr. Kawasaki, Dr. Kusumawati and Ms. Kawamura performed genotyping and statistical analyses. Dr. Kondo, Dr. Kusaoi, Dr. Amano, Dr. Kusanagi, Dr. Itoh, Dr. Fujimoto, Dr.Tamura, Dr. Hashimoto, Dr. Matsumoto and Dr. Sumida recruited the participants and collected clinical data. All authors read and approved the final version of the manuscript.

