## Supplementary material for "Genetic dissection of *HLA-DRB1*15:01* and XL9 region variants in Japanese patients with systemic lupus erythematosus: Primary role for *HLA-DRB1*15:01*": Online Supplementary Materials

Contents

Supplementary Table S1
Supplementary Table S2
Supplementary Table S3
Supplementary Table S4
Supplementary Table S5
Supplementary Table S6
Supplementary Figure S1
Supplementary Figure S2
Supplementary Data

Supplementary Table S1 Primers and probes used in this study

| | forward primer | reverse primer | VIC labeled probe | FAM labeled probe |
|---|---|---|---|---|
| rs9271375<br>rs9271378 | TGGTCTCTCTGAATCTCAACWTTT* | AATCAGCATAGTGATTSCACAATA | NA | NA |
| rs2105898 | GGCTCTTCCACACTCAYTCACAGT | CAGAGTTAGGGCCACTGTCTCAA | CCCTGGTTAATGTATTC | CCCTGGTTAATGTAGTC |
| rs9271593 | GCATCATTTGTCTCTGGCAGACT | CTCCATCCAACTTCATGATTCATT | TGTTGAGTCTATGTATTTC | TGTTGAGTCTATATATTTC |

NA, not applicable.

*Forward primer was used for amplification and cycle sequencing reaction.

Supplementary Table S2. Data on XL9 variants in GWAS of SLE in a Japanese population.

| CHR | POS | SNV | effect allele | BETA | SE | P | effect allele frequency | |
|---|---|---|---|---|---|---|---|---|
| | | | | | | | SLE | controls |
| 6 | 32587067 | rs9271375 | A | -0.320 | 0.084 | $1.4x10^{-4}$ | 0.273 | 0.346 |
| 6 | 32587300 | rs9271378 | G | -0.393 | 0.115 | $6.1x10^{-4}$ | 0.095 | 0.143 |
| 6 | 32590498 | rs2105898 | G* | -0.203 | 0.094 | 0.031 | 0.730 | 0.765 |
| 6 | 32591198 | rs9271593 | T* | -0.011 | 0.085 | 0.90 | 0.680 | 0.681 |

Based on GWAS reported by Sakaue et al. [19]

*The reported risk alleles of rs2105898 and rs9271593 are T and C, respectively.

Data was obtained from the NBDC Human Database (https://humandbs.biosciencedbc.jp/en/, Dataset ID: hum0197.v3.gwas.v1)

Supplementary Table S3. Association analysis of XL9 variants with subsets of SLE

| | rs9271375A | | | rs9271378G | | | rs2105898T | | | rs9271593C | | |
|---|---|---|---|---|---|---|---|---|---|---|---|---|
| | P | FDR Q* | OR (95% CI) | P | FDR Q* | OR (95% CI) | P | FDR Q* | OR (95% CI) | P | FDR Q* | OR (95% CI) |
| Age of onset | | | | | | | | | | | | |
| < 20 years versus ≥ 20 years | 0.75 | 0.87 | 0.94 (0.63-1.37) | 0.59 | 0.79 | 1.15 (0.67-1.91) | 0.29 | 0.63 | 1.21 (0.85-1.73) | 0.22 | 0.63 | 1.24 (0.87-1.77) |
| Renal disorder | | | | | | | | | | | | |
| Present versus Absent | 0.29 | 0.63 | 0.84 (0.62-1.16) | 0.36 | 0.63 | 1.23 (0.79-1.94) | 0.51 | 0.74 | 0.90 (0.67-1.22) | 0.42 | 0.68 | 0.89 (0.66-1.19) |
| Anti-dsDNA antibody | | | | | | | | | | | | |
| Present versus Absent | 0.87 | 0.87 | 0.97 (0.64-1.49) | 0.33 | 0.63 | 0.76 (0.45-1.35) | 0.82 | 0.87 | 1.05 (0.71-1.58) | 0.85 | 0.87 | 0.96 (0.66-1.43) |
| Anti-Sm antibody | | | | | | | | | | | | |
| Present versus Absent | 0.12 | 0.48 | 0.72 (0.46-1.08) | 0.054 | 0.44 | 0.51 (0.25-0.97) | 0.11 | 0.48 | 0.72 (0.48-1.07) | 0.011 | 0.18 | 0.60 (0.40-0.88) |

FDR, false discovery rate; OR, odds ratio; CI, confidence interval.

* Significance level was set at FDR Q<0.05.

Supplementary Table S4. Comparisons of allele frequencies and linkage disequilibrium status of *HLA-DRB1*15* and XL9 variants among European, East Asian, and African populations

| | CEU | | | JPT | | | CHB | | | YRI | | |
|---|---|---|---|---|---|---|---|---|---|---|---|---|
| | Frequency | LD ($r^2$) with | | Frequency | LD ($r^2$) with | | Frequency | LD ($r^2$) with | | Frequency | LD ($r^2$) with | |
| | (%) | rs2105898 | rs9271593 | (%) | rs2105898 | rs9271593 | (%) | rs2105898 | rs9271593 | (%) | rs2105898 | rs9271593 |
| XL9 | | | | | | | | | | | | |
| rs2105898T | 19.2 | NA | 0.497 | 28.8 | NA | 0.725 | 21.4 | NA | 0428 | 19.9 | NA | 0.210 |
| rs9271593C | 32.3 | 0.497 | NA | 34.6 | 0.725 | NA | 38.8 | 0.428 | NA | 54.2 | 0.210 | NA |
| *HLA-DRB1* | | | | | | | | | | | | |
| **15:01* | 16.2 | 0.782 | 0.389 | 8.1 | 0.220 | 0.168 | 9.3 | 0.396 | 0.169 | 0.5 | 0.019 | 0.005 |
| **15:02* | 0.5 | 0.021 | 0.011 | 12.9 | 0.352 | 0.270 | 3.7 | 0.130 | 0.055 | 0 | NA | NA |
| **15:03* | 0 | NA | NA | 0 | NA | NA | 0 | NA | NA | 15.3 | 0.674 | 0.142 |

CEU, Utah residents (CEPH) with Northern and Western European ancestry from the CEPH collection; JPT, Japanese in Tokyo, Japan; CHB, Han Chinese in Beijing, China; YRI, Yoruba in Ibadan, Nigeria; LD, linkage disequilibrium; NA, not applicable.

Genotype data of *HLA-DRB1* alleles, rs2105898 and rs9271593 in the CEU, JPT, CHB, and YRI were obtained from the International Genome Sample Resource (2018 data, https://www.internationalgenome.org/category/hla/, accessed 28 Jun 2022) and the Ensembl database (Release 106, https://asia.ensembl.org/index.html, accessed 28 Jun 2022)[20].

$r^2$ values were calculated by the Haploview software.

Supplementary Table S5. Pair-wise linkage equilibrium values ($r^2$) between *HLA-DRB1* allele and XL9 region SNVs in the Japanese population.

| *HLA-DRB1* | rs2105898 | rs9271593 | rs9271375 | rs9271378 |
|---|---|---|---|---|
| 01:01 | 0.021 | 0.031 | 0.03 | 0.393 |
| 03:01 | 0 | 0.003 | 0.003 | 0.008 |
| 04:01 | 0.004 | 0.006 | 0.026 | 0.002 |
| 04:03 | 0.009 | 0.014 | 0.061 | 0.005 |
| 04:04 | 0.001 | 0.001 | 0.004 | 0 |
| 04:05 | 0.043 | 0.058 | 0.272 | 0.016 |
| 04:06 | 0.011 | 0.017 | 0.075 | 0.003 |
| 04:07 | 0.001 | 0 | 0.011 | 0.001 |
| 04:10 | 0.005 | 0.008 | 0.035 | 0.003 |
| 07:01 | 0 | 0 | 0.002 | 0.02 |
| 08:02 | 0.014 | 0.003 | 0.005 | 0.007 |
| 08:03 | 0.018 | 0.033 | 0.039 | 0.013 |
| 09:01 | 0.054 | 0.079 | 0.074 | 0.027 |
| 10:01 | 0 | 0 | 0.001 | 0 |
| 11:01 | 0.006 | 0.036 | 0.036 | 0.11 |
| 12:01 | 0.012 | 0.017 | 0.02 | 0.06 |
| 12:02 | 0.049 | 0.037 | 0.01 | 0.001 |
| 13:01 | 0.002 | 0 | 0.005 | 0 |
| 13:02 | 0.029 | 0.042 | 0.036 | 0.014 |
| 14:03 | 0.007 | 0.044 | 0.044 | 0.128 |
| 14:05 | 0.01 | 0.015 | 0.014 | 0.005 |
| 14:06 | 0.003 | 0.022 | 0.022 | 0.063 |
| 14:54 | 0.011 | 0.016 | 0.016 | 0.005 |
| 15:01 | 0.213 | 0.15 | 0.03 | 0.012 |
| 15:02 | 0.431 | 0.288 | 0.066 | 0.023 |
| 16:02 | 0.025 | 0.016 | 0.005 | 0.002 |

Linkage disequilibrium values ($r^2$) were calculated using Haploview software based on the genotypes of 779 healthy Japanese individuals. LD plot for *DRB1*15:0*1 and *DRB1*15:02* is shown in Supplementary Figure S2.

Supplementary Table S6. Association of XL9 variants with the expression of *HLA* genes in the GTEx Portal

| Gene | XL9 variant | P | Effect allele | NES | SLE risk allele | Effect of the risk allele | Tissue |
|---|---|---|---|---|---|---|---|
| *HLA-DRB9* | rs2105898 | $3.9 \times 10^{-32}$ | G | 0.60 | T | down | Whole Blood |
| *HLA-DRB5* | rs2105898 | $3.1 \times 10^{-94}$ | G | -0.78 | T | up | Whole Blood |
| *HLA-DRB6* | rs2105898 | $2.3 \times 10^{-59}$ | G | 0.80 | T | down | Whole Blood |
| *HLA-DRB1* | rs2105898 | $4.7 \times 10^{-6}$ | G | -0.10 | T | up | Whole Blood |
| *HLA-DQA1* | rs2105898 | $3.4 \times 10^{-8}$ | G | -0.15 | T | up | Whole Blood |
| *HLA-DQB1* | rs2105898 | $6.6 \times 10^{-20}$ | G | -0.39 | T | up | Whole Blood |
| *HLA-DQA2* | rs2105898 | $3.2 \times 10^{-15}$ | G | 0.49 | T | down | Whole Blood |
| *HLA-DQB2* | rs2105898 | $2.1 \times 10^{-12}$ | G | 0.39 | T | down | Whole Blood |
| *HLA-DRB9* | rs9271593 | $8.7 \times 10^{-10}$ | T | 0.25 | C | down | Whole Blood |
| *HLA-DRB5* | rs9271593 | $3.9 \times 10^{-13}$ | T | -0.24 | C | up | Whole Blood |
| *HLA-DRB6* | rs9271593 | $4.1 \times 10^{-46}$ | T | 0.55 | C | down | Whole Blood |
| *HLA-DRB1* | rs9271593 | $9.7 \times 10^{-17}$ | T | -0.14 | C | up | Whole Blood |
| *HLA-DQA2* | rs9271593 | $1.4 \times 10^{-15}$ | T | 0.38 | C | down | Whole Blood |

NES, normalized effect size.

eQTL data of XL9 rs2105898 and rs9271593 in whole blood was obtained from the Genotype-Tissue Expression (GTEx) Portal V8 (https://gtexportal.org/home/, accessed 13 Mar 2023).

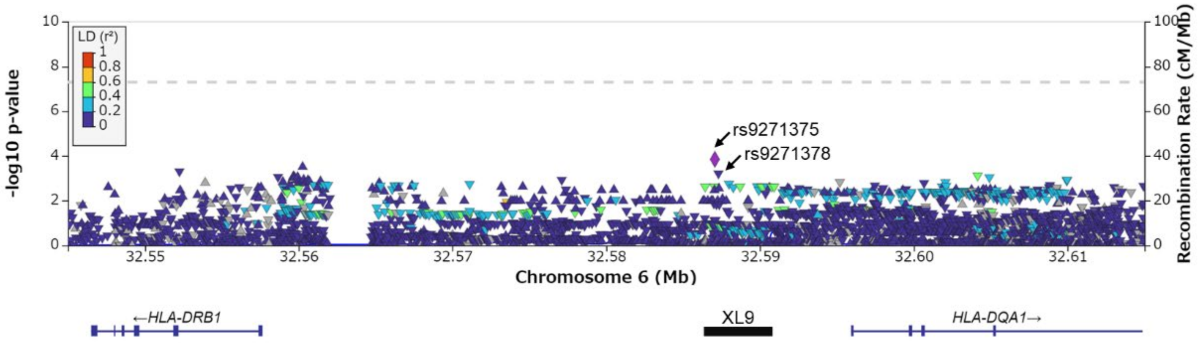


**Supplementary Figure S1. Regional plot of XL9 variants in Genome-wide association study of Systemic lupus erythematosus in a Japanese population.** GWAS data was obtained from the NBDC Human Database (Dataset ID: hum0197.v3.gwas.v1).[19] Regional plot of a region between *HLA-DRB1* and *DQA1* was drawn by LocusZoom (http://locuszoom.org/). Color of dots shows level of linkage disequilibrium between rs9271375 and each variant in the East Asian populations.

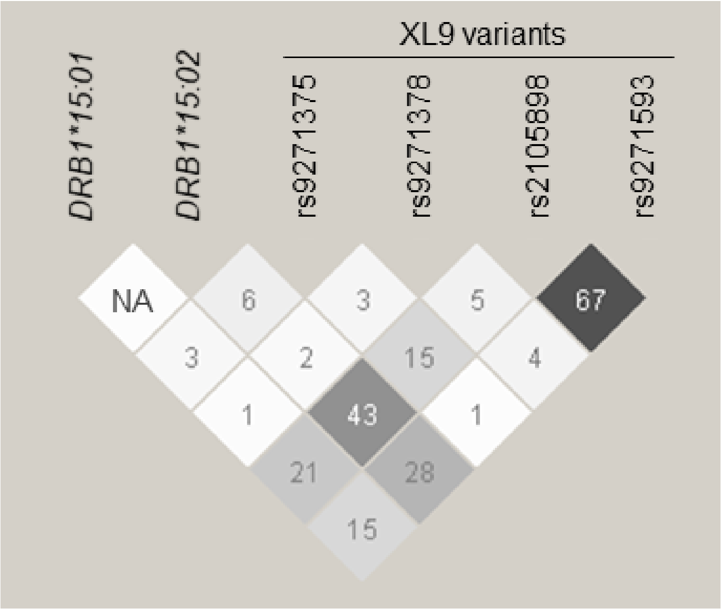


**Supplementary Figure S2. Linkage disequilibrium among the XL9 region variants and *HLA-DRB1* alleles examined in this study.**

Pair-wise linkage equilibrium values ($r^2$) were calculated using Haploview software based on the genotypes of 779 healthy individuals. Data for all alleles are is shown in Supplementary Table S5. NA: not applicable.

Supplementary Data Genotyping data of sach subject

| ID | DRB1*15:01 allele count | DRB1*15:02 allele count | rs9271375 genotype | rs9271375A allele count | rs9271378 genotype | rs9271378G allele count | rs2105898 genotype | rs2105898T allele count | rs9271593 genotype | rs9271593C allele count |
|---|---|---|---|---|---|---|---|---|---|---|
| SLE1 | 0 | 0 | G/G | 0 | G/A | 1 | G/G | | T/T | 0 |
| SLE2 | 0 | 0 | A/G | 1 | A/A | 0 | T/G | 1 | C/T | 1 |
| SLE3 | 0 | 1 | G/G | 0 | A/A | 0 | T/G | 1 | C/T | 1 |
| SLE4 | 1 | 0 | G/G | 0 | A/A | 0 | T/G | 1 | C/T | 1 |
| SLE5 | 0 | 2 | G/G | 0 | A/A | 0 | T/T | 2 | C/C | 2 |
| SLE6 | 0 | 0 | G/G | 0 | G/A | 1 | G/G | 0 | T/T | 0 |
| SLE7 | 0 | 1 | A/G | 1 | G/A | 1 | G/G | 0 | T/T | 0 |
| SLE8 | 1 | 0 | G/G | 0 | A/A | 0 | T/G | 1 | C/T | 1 |
| SLE9 | 0 | 0 | G/G | 0 | A/A | 0 | G/G | 0 | T/T | 0 |
| SLE10 | 0 | 0 | G/G | 0 | G/A | 1 | G/G | 0 | T/T | 0 |
| SLE11 | 1 | 0 | A/G | 1 | G/A | 1 | T/G | 1 | C/C | 2 |
| SLE12 | 0 | 0 | A/G | 1 | G/A | 1 | G/G | 0 | C/T | 1 |
| SLE13 | 0 | 0 | A/G | 1 | G/A | 1 | G/G | 0 | C/T | 1 |
| SLE14 | 0 | 0 | A/G | 1 | A/A | 0 | G/G | 0 | T/T | 0 |
| SLE15 | 0 | 1 | G/G | 0 | A/A | 0 | T/G | 1 | C/T | 1 |
| SLE16 | 0 | 0 | A/G | 1 | A/A | 0 | G/G | 0 | T/T | 0 |
| SLE17 | 0 | 0 | G/G | 0 | A/A | 0 | G/G | 0 | T/T | 0 |
| SLE18 | 0 | 0 | G/G | 0 | G/A | 1 | G/G | 0 | T/T | 0 |
| SLE19 | 0 | 1 | G/G | 0 | G/A | 1 | T/G | 1 | C/T | 1 |
| SLE20 | 0 | 1 | G/G | 0 | A/A | 0 | T/G | 1 | C/T | 1 |
| SLE21 | 0 | 0 | G/G | 0 | A/A | 0 | G/G | 0 | T/T | 0 |
| SLE22 | 2 | 0 | G/G | 0 | A/A | 0 | T/T | 2 | C/C | 2 |
| SLE23 | 1 | 0 | G/G | 0 | A/A | 0 | T/T | 2 | C/C | 2 |
| SLE24 | 2 | 0 | G/G | 0 | A/A | 0 | T/T | 2 | C/C | 2 |
| SLE25 | 0 | 0 | A/G | 1 | A/A | 0 | G/G | 0 | T/T | 0 |
| SLE26 | 2 | 0 | G/G | 0 | A/A | 0 | T/T | 2 | C/C | 2 |
| SLE27 | 0 | 1 | A/G | 1 | A/A | 0 | T/G | 1 | C/T | 1 |
| SLE28 | 1 | 0 | A/G | 1 | A/A | 0 | T/G | 1 | C/T | 1 |
| SLE29 | 0 | 0 | G/G | 0 | A/A | 0 | G/G | 0 | T/T | 0 |
| SLE30 | 0 | 0 | G/G | 0 | A/A | 0 | T/G | 1 | C/T | 1 |
| SLE31 | 1 | 0 | G/G | 0 | A/A | 0 | T/T | 2 | C/C | 2 |
| SLE32 | 0 | 0 | A/A | 2 | A/A | 0 | G/G | 0 | T/T | 0 |
| SLE33 | 2 | 0 | G/G | 0 | G/A | 1 | T/G | 1 | C/T | 1 |
| SLE34 | 0 | 0 | A/G | 1 | G/G | 2 | G/G | 0 | C/T | 1 |
| SLE35 | 0 | 0 | A/G | 1 | A/A | 0 | G/G | 0 | T/T | 0 |
| SLE36 | 0 | 0 | A/A | 2 | A/A | 0 | G/G | 0 | T/T | 0 |
| SLE37 | 1 | 0 | G/G | 0 | A/A | 0 | T/G | 1 | C/T | 1 |
| SLE38 | 0 | 0 | A/A | 2 | A/A | 0 | G/G | 0 | T/T | 0 |
| SLE39 | 1 | 0 | A/G | 1 | A/A | 0 | T/G | 1 | C/T | 1 |
| SLE40 | 0 | 1 | G/G | 0 | G/A | 1 | T/G | 1 | C/T | 1 |
| SLE41 | 0 | 0 | A/G | 1 | A/A | 0 | G/G | 0 | T/T | 0 |
| SLE42 | 0 | 0 | A/A | 2 | G/G | 2 | G/G | 0 | C/C | 2 |
| SLE43 | 0 | 0 | A/G | 1 | A/A | 0 | G/G | 0 | T/T | 0 |
| SLE44 | 1 | 0 | G/G | 0 | A/A | 0 | T/G | 1 | C/T | 1 |

| | | | | | | | | | | |
|---|---|---|---|---|---|---|---|---|---|---|
| SLE45 | 0 | 0 | A/G | 1 | A/A | 0 | G/G | 0 | T/T | 0 |
| SLE46 | 1 | 0 | G/G | 0 | A/A | 0 | T/G | 1 | C/T | 1 |
| SLE47 | 0 | 2 | G/G | 0 | A/A | 0 | T/T | 2 | C/C | 2 |
| SLE48 | 1 | 0 | G/G | 0 | A/A | 0 | T/G | 1 | C/T | 1 |
| SLE49 | 0 | 0 | A/A | 2 | G/G | 2 | G/G | 0 | C/C | 2 |
| SLE50 | 0 | 2 | G/G | 0 | A/A | 0 | T/G | 1 | C/T | 1 |
| SLE51 | 0 | 0 | A/G | 1 | A/A | 0 | G/G | 0 | T/T | 0 |
| SLE52 | 1 | 0 | G/G | 0 | A/A | 0 | T/T | 2 | C/C | 2 |
| SLE53 | 0 | 0 | A/A | 2 | G/A | 1 | G/G | 0 | C/T | 1 |
| SLE54 | 0 | 1 | A/G | 1 | A/A | 0 | T/G | 1 | C/T | 1 |
| SLE55 | 0 | 0 | A/A | 2 | G/A | 1 | G/G | 0 | C/T | 1 |
| SLE56 | 1 | 0 | G/G | 0 | A/A | 0 | T/T | 2 | C/C | 2 |
| SLE57 | 0 | 0 | A/G | 1 | G/A | 1 | G/G | 0 | C/T | 1 |
| SLE58 | 0 | 0 | G/G | 0 | A/A | 0 | G/G | 0 | T/T | 0 |
| SLE59 | 0 | 0 | G/G | 0 | A/A | 0 | G/G | 0 | T/T | 0 |
| SLE60 | 1 | 0 | G/G | 0 | A/A | 0 | T/G | 1 | C/T | 1 |
| SLE61 | 0 | 0 | A/G | 1 | A/A | 0 | G/G | 0 | T/T | 0 |
| SLE62 | 0 | 0 | G/G | 0 | A/A | 0 | G/G | 0 | T/T | 0 |
| SLE63 | 0 | 0 | G/G | 0 | G/A | 1 | G/G | 0 | T/T | 0 |
| SLE64 | 0 | 1 | G/G | 0 | A/A | 0 | T/G | 1 | C/T | 1 |
| SLE65 | 0 | 0 | A/G | 1 | G/A | 1 | G/G | 0 | C/T | 1 |
| SLE66 | 0 | 0 | A/G | 1 | G/A | 1 | G/G | 0 | T/T | 0 |
| SLE67 | 0 | 1 | A/G | 1 | A/A | 0 | T/G | 1 | C/T | 1 |
| SLE68 | 0 | 0 | G/G | 0 | A/A | 0 | G/G | 0 | T/T | 0 |
| SLE69 | 0 | 0 | A/G | 1 | G/A | 1 | G/G | 0 | T/T | 0 |
| SLE70 | 0 | 1 | G/G | 0 | A/A | 0 | T/G | 1 | C/T | 1 |
| SLE71 | 0 | 0 | A/G | 1 | G/A | 1 | G/G | 0 | C/T | 1 |
| SLE72 | 0 | 0 | G/G | 0 | G/A | 1 | G/G | 0 | T/T | 0 |
| SLE73 | 0 | 0 | A/G | 1 | G/A | 1 | G/G | 0 | C/T | 1 |
| SLE74 | 1 | 0 | G/G | 0 | A/A | 0 | T/G | 1 | C/T | 1 |
| SLE75 | 2 | 0 | G/G | 0 | A/A | 0 | T/T | 2 | C/C | 2 |
| SLE76 | 0 | 0 | G/G | 0 | A/A | 0 | G/G | 0 | T/T | 0 |
| SLE77 | 0 | 1 | A/G | 1 | A/A | 0 | T/G | 1 | C/T | 1 |
| SLE78 | 1 | 0 | G/G | 0 | A/A | 0 | T/G | 1 | C/T | 1 |
| SLE79 | 1 | 0 | A/G | 1 | A/A | 0 | T/G | 1 | C/T | 1 |
| SLE80 | 0 | 0 | A/G | 1 | G/A | 1 | T/G | 1 | C/C | 2 |
| SLE81 | 0 | 0 | G/G | 0 | A/A | 0 | G/G | 0 | T/T | 0 |
| SLE82 | 0 | 0 | A/G | 1 | A/A | 0 | G/G | 0 | T/T | 0 |
| SLE83 | 0 | 0 | A/G | 1 | A/A | 0 | G/G | 0 | T/T | 0 |
| SLE84 | 0 | 0 | G/G | 0 | A/A | 0 | G/G | 0 | T/T | 0 |
| SLE85 | 0 | 0 | A/G | 1 | G/A | 1 | G/G | 0 | T/T | 0 |
| SLE86 | 1 | 0 | G/G | 0 | A/A | 0 | T/G | 1 | C/T | 1 |
| SLE87 | 0 | 0 | G/G | 0 | A/A | 0 | G/G | 0 | T/T | 0 |
| SLE88 | 0 | 0 | G/G | 0 | A/A | 0 | G/G | 0 | T/T | 0 |
| SLE89 | 0 | 0 | G/G | 0 | A/A | 0 | T/G | 1 | C/T | 1 |
| SLE90 | 0 | 1 | G/G | 0 | A/A | 0 | T/G | 1 | C/T | 1 |
| SLE91 | 0 | 0 | G/G | 0 | A/A | 0 | G/G | 0 | T/T | 0 |
| SLE92 | 0 | 0 | A/A | 2 | G/A | 1 | G/G | 0 | C/T | 1 |
| SLE93 | 0 | 1 | G/G | 0 | A/A | 0 | T/G | 1 | C/T | 1 |

| | | | | | | | | | | |
|---|---|---|---|---|---|---|---|---|---|---|
| SLE94 | 1 | 0 | A/G | 1 | A/A | 0 | T/G | 1 | C/T | 1 |
| SLE95 | 0 | 0 | A/A | 2 | A/A | 0 | G/G | 0 | T/T | 0 |
| SLE96 | 0 | 1 | A/G | 1 | A/A | 0 | T/G | 1 | C/T | 1 |
| SLE97 | 0 | 1 | G/G | 0 | A/A | 0 | T/G | 1 | C/T | 1 |
| SLE98 | 0 | 0 | A/G | 1 | A/A | 0 | G/G | 0 | T/T | 0 |
| SLE99 | 0 | 0 | A/G | 1 | A/A | 0 | G/G | 0 | T/T | 0 |
| SLE100 | 0 | 0 | G/G | 0 | A/A | 0 | G/G | 0 | T/T | 0 |
| SLE101 | 0 | 0 | G/G | 0 | A/A | 0 | T/G | 1 | C/T | 1 |
| SLE102 | 0 | 0 | A/G | 1 | G/A | 1 | G/G | 0 | C/T | 1 |
| SLE103 | 0 | 0 | A/G | 1 | A/A | 0 | G/G | 0 | T/T | 0 |
| SLE104 | 0 | 1 | A/G | 1 | A/A | 0 | T/G | 1 | C/T | 1 |
| SLE105 | 0 | 0 | G/G | 0 | A/A | 0 | G/G | 0 | T/T | 0 |
| SLE106 | 0 | 0 | G/G | 0 | A/A | 0 | T/T | 2 | C/C | 2 |
| SLE107 | 1 | 0 | G/G | 0 | A/A | 0 | T/T | 2 | C/C | 2 |
| SLE108 | 1 | 0 | G/G | 0 | A/A | 0 | T/G | 1 | C/T | 1 |
| SLE109 | 0 | 1 | A/G | 1 | G/A | 1 | T/G | 1 | C/C | 2 |
| SLE110 | 1 | 0 | A/G | 1 | A/A | 0 | T/G | 1 | C/T | 1 |
| SLE111 | 1 | 0 | G/G | 0 | A/A | 0 | T/G | 1 | C/T | 1 |
| SLE112 | 0 | 1 | A/G | 1 | G/A | 1 | T/G | 1 | C/C | 2 |
| SLE113 | 0 | 2 | G/G | 0 | A/A | 0 | T/T | 2 | C/C | 2 |
| SLE114 | 0 | 0 | G/G | 0 | G/A | 1 | G/G | 0 | T/T | 0 |
| SLE115 | 2 | 0 | G/G | 0 | A/A | 0 | T/T | 2 | C/C | 2 |
| SLE116 | 0 | 0 | G/G | 0 | A/A | 0 | G/G | 0 | T/T | 0 |
| SLE117 | 1 | 0 | G/G | 0 | A/A | 0 | T/G | 1 | C/T | 1 |
| SLE118 | 0 | 0 | A/G | 1 | A/A | 0 | G/G | 0 | T/T | 0 |
| SLE119 | 1 | 0 | A/G | 1 | G/A | 1 | T/G | 1 | C/C | 2 |
| SLE120 | 0 | 0 | A/G | 1 | G/A | 1 | G/G | 0 | C/T | 1 |
| SLE121 | 0 | 0 | A/G | 1 | A/A | 0 | G/G | 0 | T/T | 0 |
| SLE122 | 0 | 1 | A/G | 1 | A/A | 0 | T/G | 1 | C/T | 1 |
| SLE123 | 1 | 0 | G/G | 0 | A/A | 0 | T/G | 1 | C/T | 1 |
| SLE124 | 1 | 0 | A/G | 1 | A/A | 0 | T/G | 1 | C/T | 1 |
| SLE125 | 1 | 0 | G/G | 0 | A/A | 0 | T/G | 1 | C/T | 1 |
| SLE126 | 0 | 0 | G/G | 0 | A/A | 0 | G/G | 0 | T/T | 0 |
| SLE127 | 0 | 0 | G/G | 0 | A/A | 0 | G/G | 0 | T/T | 0 |
| SLE128 | 0 | 0 | G/G | 0 | G/A | 1 | G/G | 0 | T/T | 0 |
| SLE129 | 0 | 0 | A/G | 1 | A/A | 0 | G/G | 0 | T/T | 0 |
| SLE130 | 0 | 0 | A/G | 1 | A/A | 0 | G/G | 0 | T/T | 0 |
| SLE131 | 0 | 0 | A/A | 2 | G/A | 1 | G/G | 0 | C/T | 1 |
| SLE132 | 0 | 0 | A/A | 2 | G/A | 1 | G/G | 0 | C/T | 1 |
| SLE133 | 0 | 0 | G/G | 0 | A/A | 0 | G/G | 0 | T/T | 0 |
| SLE134 | 0 | 0 | G/G | 0 | G/A | 1 | G/G | 0 | T/T | 0 |
| SLE135 | 0 | 0 | G/G | 0 | A/A | 0 | G/G | 0 | T/T | 0 |
| SLE136 | 0 | 1 | A/G | 1 | A/A | 0 | T/G | 1 | C/T | 1 |
| SLE137 | 0 | 0 | G/G | 0 | G/A | 1 | G/G | 0 | T/T | 0 |
| SLE138 | 0 | 0 | A/G | 1 | A/A | 0 | G/G | 0 | T/T | 0 |
| SLE139 | 0 | 0 | A/G | 1 | A/A | 0 | G/G | 0 | T/T | 0 |
| SLE140 | 0 | 0 | G/G | 0 | A/A | 0 | G/G | 0 | T/T | 0 |
| SLE141 | 1 | 0 | G/G | 0 | A/A | 0 | T/G | 1 | C/T | 1 |
| SLE142 | 0 | 0 | A/G | 1 | A/A | 0 | G/G | 0 | T/T | 0 |

| | | | | | | | | | | |
|---|---|---|---|---|---|---|---|---|---|---|
| SLE143 | 0 | 0 | A/A | 2 | G/A | 1 | G/G | 0 | C/T | 1 |
| SLE144 | 0 | 0 | A/G | 1 | A/A | 0 | G/G | 0 | T/T | 0 |
| SLE145 | 0 | 0 | A/G | 1 | A/A | 0 | T/G | 1 | C/T | 1 |
| SLE146 | 0 | 0 | G/G | 0 | A/A | 0 | G/G | 0 | T/T | 0 |
| SLE147 | 1 | 0 | G/G | 0 | A/A | 0 | T/G | 1 | C/T | 1 |
| SLE148 | 0 | 0 | G/G | 0 | A/A | 0 | G/G | 0 | T/T | 0 |
| SLE149 | 0 | 0 | A/G | 1 | A/A | 0 | G/G | 0 | T/T | 0 |
| SLE150 | 0 | 0 | A/A | 2 | G/A | 1 | G/G | 0 | C/T | 1 |
| SLE151 | 0 | 0 | A/G | 1 | A/A | 0 | G/G | 0 | T/T | 0 |
| SLE152 | 0 | 0 | A/G | 1 | A/A | 0 | G/G | 0 | T/T | 0 |
| SLE153 | 0 | 0 | G/G | 0 | A/A | 0 | G/G | 0 | T/T | 0 |
| SLE154 | 0 | 0 | A/G | 1 | G/A | 1 | G/G | 0 | C/T | 1 |
| SLE155 | 1 | 0 | G/G | 0 | A/A | 0 | T/T | 2 | C/C | 2 |
| SLE156 | 1 | 0 | G/G | 0 | A/A | 0 | T/G | 1 | C/T | 1 |
| SLE157 | 1 | 0 | G/G | 0 | A/A | 0 | T/T | 2 | C/C | 2 |
| SLE158 | 0 | 1 | A/G | 1 | A/A | 0 | T/G | 1 | C/T | 1 |
| SLE159 | 0 | 0 | G/G | 0 | G/A | 1 | G/G | 0 | T/T | 0 |
| SLE160 | 1 | 1 | G/G | 0 | A/A | 0 | T/T | 2 | C/C | 2 |
| SLE161 | 1 | 0 | G/G | 0 | A/A | 0 | T/G | 1 | C/T | 1 |
| SLE162 | 0 | 0 | A/G | 1 | G/A | 1 | G/G | 0 | C/T | 1 |
| SLE163 | 0 | 0 | G/G | 0 | A/A | 0 | T/G | 1 | C/T | 1 |
| SLE164 | 0 | 0 | A/G | 1 | A/A | 0 | G/G | 0 | T/T | 0 |
| SLE165 | 0 | 0 | G/G | 0 | A/A | 0 | G/G | 0 | T/T | 0 |
| SLE166 | 0 | 0 | A/G | 1 | G/G | 2 | G/G | 0 | C/T | 1 |
| SLE167 | 1 | 0 | A/G | 1 | A/A | 0 | T/G | 1 | C/T | 1 |
| SLE168 | 1 | 0 | G/G | 0 | A/A | 0 | T/G | 1 | C/T | 1 |
| SLE169 | 1 | 0 | A/G | 1 | A/A | 0 | T/G | 1 | C/T | 1 |
| SLE170 | 0 | 0 | A/G | 1 | A/A | 0 | G/G | 0 | T/T | 0 |
| SLE171 | 0 | 0 | G/G | 0 | A/A | 0 | G/G | 0 | T/T | 0 |
| SLE172 | 1 | 0 | G/G | 0 | G/A | 1 | T/G | 1 | C/T | 1 |
| SLE173 | 0 | 0 | A/G | 1 | G/A | 1 | G/G | 0 | C/T | 1 |
| SLE174 | 0 | 1 | A/G | 1 | G/A | 1 | T/G | 1 | C/C | 2 |
| SLE175 | 0 | 0 | G/G | 0 | A/A | 0 | G/G | 0 | T/T | 0 |
| SLE176 | 0 | 0 | A/G | 1 | A/A | 0 | G/G | 0 | T/T | 0 |
| SLE177 | 0 | 0 | G/G | 0 | A/A | 0 | G/G | 0 | T/T | 0 |
| SLE178 | 0 | 0 | A/G | 1 | G/A | 1 | G/G | 0 | T/T | 0 |
| SLE179 | 0 | 0 | G/G | 0 | A/A | 0 | G/G | 0 | T/T | 0 |
| SLE180 | 0 | 0 | A/G | 1 | A/A | 0 | G/G | 0 | T/T | 0 |
| SLE181 | 0 | 1 | G/G | 0 | A/A | 0 | T/G | 1 | C/T | 1 |
| SLE182 | 1 | 0 | G/G | 0 | A/A | 0 | T/G | 1 | C/T | 1 |
| SLE183 | 0 | 1 | A/G | 1 | A/A | 0 | T/G | 1 | C/T | 1 |
| SLE184 | 0 | 1 | A/G | 1 | A/A | 0 | T/G | 1 | C/T | 1 |
| SLE185 | 1 | 0 | G/G | 0 | A/A | 0 | T/G | 1 | C/T | 1 |
| SLE186 | 0 | 0 | G/G | 0 | A/A | 0 | G/G | 0 | T/T | 0 |
| SLE187 | 0 | 0 | A/G | 1 | A/A | 0 | G/G | 0 | T/T | 0 |
| SLE188 | 0 | 1 | G/G | 0 | A/A | 0 | T/G | 1 | C/T | 1 |
| SLE189 | 0 | 0 | A/A | 2 | G/A | 1 | G/G | 0 | C/T | 1 |
| SLE190 | 0 | 1 | G/G | 0 | A/A | 0 | T/G | 1 | C/T | 1 |
| SLE191 | 0 | 0 | G/G | 0 | A/A | 0 | T/T | 2 | C/C | 2 |

| | | | | | | | | | | |
|---|---|---|---|---|---|---|---|---|---|---|
| SLE192 | 2 | 0 | G/G | 0 | A/A | 0 | T/T | 2 | C/C | 2 |
| SLE193 | 0 | 0 | G/G | 0 | G/A | 1 | G/G | 0 | T/T | 0 |
| SLE194 | 1 | 0 | G/G | 0 | A/A | 0 | T/G | 1 | C/T | 1 |
| SLE195 | 0 | 0 | A/A | 2 | G/A | 1 | G/G | 0 | C/T | 1 |
| SLE196 | 0 | 1 | G/G | 0 | A/A | 0 | T/T | 2 | C/C | 2 |
| SLE197 | 0 | 0 | A/A | 2 | G/A | 1 | G/G | 0 | C/T | 1 |
| SLE198 | 0 | 0 | G/G | 0 | A/A | 0 | G/G | 0 | T/T | 0 |
| SLE199 | 0 | 0 | A/G | 1 | A/A | 0 | G/G | 0 | T/T | 0 |
| SLE200 | 0 | 0 | G/G | 0 | A/A | 0 | T/G | 1 | C/T | 1 |
| SLE201 | 0 | 0 | A/A | 2 | A/A | 0 | G/G | 0 | T/T | 0 |
| SLE202 | 1 | 1 | G/G | 0 | A/A | 0 | T/T | 2 | C/C | 2 |
| SLE203 | 0 | 0 | A/G | 1 | A/A | 0 | G/G | 0 | T/T | 0 |
| SLE204 | 0 | 0 | A/G | 1 | A/A | 0 | G/G | 0 | T/T | 0 |
| SLE205 | 0 | 1 | G/G | 0 | A/A | 0 | T/G | 1 | C/T | 1 |
| SLE206 | 0 | 0 | A/G | 1 | A/A | 0 | T/G | 1 | C/T | 1 |
| SLE207 | 0 | 0 | G/G | 0 | G/A | 1 | T/G | 1 | C/T | 1 |
| SLE208 | 0 | 0 | A/G | 1 | A/A | 0 | T/G | 1 | C/T | 1 |
| SLE209 | 0 | 0 | G/G | 0 | A/A | 0 | G/G | 0 | T/T | 0 |
| SLE210 | 1 | 0 | A/G | 1 | A/A | 0 | T/G | 1 | C/T | 1 |
| SLE211 | 0 | 0 | G/G | 0 | A/A | 0 | G/G | 0 | T/T | 0 |
| SLE212 | 0 | 1 | A/G | 1 | A/A | 0 | T/G | 1 | C/T | 1 |
| SLE213 | 0 | 1 | A/G | 1 | A/A | 0 | T/G | 1 | C/T | 1 |
| SLE214 | 1 | 0 | G/G | 0 | A/A | 0 | T/G | 1 | C/T | 1 |
| SLE215 | 0 | 0 | A/A | 2 | A/A | 0 | G/G | 0 | T/T | 0 |
| SLE216 | 0 | 0 | G/G | 0 | A/A | 0 | G/G | 0 | T/T | 0 |
| SLE217 | 0 | 0 | G/G | 0 | A/A | 0 | G/G | 0 | T/T | 0 |
| SLE218 | 0 | 0 | A/G | 1 | A/A | 0 | G/G | 0 | T/T | 0 |
| SLE219 | 0 | 0 | A/G | 1 | A/A | 0 | T/G | 1 | C/T | 1 |
| SLE220 | 1 | 1 | G/G | 0 | A/A | 0 | T/T | 2 | C/C | 2 |
| SLE221 | 0 | 1 | G/G | 0 | A/A | 0 | T/G | 1 | C/T | 1 |
| SLE222 | 1 | 1 | G/G | 0 | A/A | 0 | T/T | 2 | C/C | 2 |
| SLE223 | 0 | 0 | G/G | 0 | A/A | 0 | T/G | 1 | C/T | 1 |
| SLE224 | 1 | 0 | G/G | 0 | A/A | 0 | T/T | 2 | C/C | 2 |
| SLE225 | 0 | 0 | A/G | 1 | G/A | 1 | T/G | 1 | C/C | 2 |
| SLE226 | 0 | 0 | A/G | 1 | G/A | 1 | G/G | 0 | T/T | 0 |
| SLE227 | 0 | 0 | G/G | 0 | A/A | 0 | G/G | 0 | T/T | 0 |
| SLE228 | 0 | 0 | A/G | 1 | G/A | 1 | G/G | 0 | C/T | 1 |
| SLE229 | 0 | 0 | A/A | 2 | A/A | 0 | G/G | 0 | T/T | 0 |
| SLE230 | 0 | 0 | A/G | 1 | A/A | 0 | G/G | 0 | T/T | 0 |
| SLE231 | 0 | 0 | G/G | 0 | A/A | 0 | G/G | 0 | T/T | 0 |
| SLE232 | 0 | 0 | A/G | 1 | G/A | 1 | G/G | 0 | T/T | 0 |
| SLE233 | 0 | 0 | G/G | 0 | A/A | 0 | T/G | 1 | C/T | 1 |
| SLE234 | 0 | 0 | A/G | 1 | G/A | 1 | G/G | 0 | C/T | 1 |
| SLE235 | 0 | 0 | A/G | 1 | G/A | 1 | G/G | 0 | T/T | 0 |
| SLE236 | 1 | 0 | G/G | 0 | A/A | 0 | T/T | 2 | C/C | 2 |
| SLE237 | 0 | 0 | G/G | 0 | A/A | 0 | T/T | 2 | C/C | 2 |
| SLE238 | 0 | 1 | A/G | 1 | A/A | 0 | T/G | 1 | C/T | 1 |
| SLE239 | 1 | 0 | A/G | 1 | A/A | 0 | T/G | 1 | C/T | 1 |
| SLE240 | 0 | 0 | A/G | 1 | G/A | 1 | G/G | 0 | C/T | 1 |

| | | | | | | | | | | |
|---|---|---|---|---|---|---|---|---|---|---|
| SLE241 | 0 | 0 | G/G | 0 | A/A | 0 | T/G | 1 | C/T | 1 |
| SLE242 | 0 | 0 | A/A | 2 | G/G | 2 | G/G | 0 | C/C | 2 |
| SLE243 | 1 | 0 | G/G | 0 | A/A | 0 | T/G | 1 | C/T | 1 |
| SLE244 | 1 | 0 | G/G | 0 | A/A | 0 | T/T | 2 | C/C | 2 |
| SLE245 | 0 | 0 | A/G | 1 | G/A | 1 | G/G | 0 | C/T | 1 |
| SLE246 | 1 | 0 | G/G | 0 | A/A | 0 | T/G | 1 | C/T | 1 |
| SLE247 | 0 | 0 | G/G | 0 | A/A | 0 | G/G | 0 | T/T | 0 |
| SLE248 | 0 | 0 | A/G | 1 | A/A | 0 | G/G | 0 | T/T | 0 |
| SLE249 | 0 | 0 | G/G | 0 | A/A | 0 | G/G | 0 | T/T | 0 |
| SLE250 | 0 | 0 | A/G | 1 | G/A | 1 | G/G | 0 | T/T | 0 |
| SLE251 | 0 | 1 | G/G | 0 | A/A | 0 | T/G | 1 | C/T | 1 |
| SLE252 | 0 | 0 | G/G | 0 | A/A | 0 | G/G | 0 | T/T | 0 |
| SLE253 | 2 | 0 | G/G | 0 | A/A | 0 | T/T | 2 | C/C | 2 |
| SLE254 | 0 | 1 | G/G | 0 | A/A | 0 | T/G | 1 | C/T | 1 |
| SLE255 | 0 | 0 | A/G | 1 | G/A | 1 | G/G | 0 | C/T | 1 |
| SLE256 | 0 | 1 | G/G | 0 | G/A | 1 | T/G | 1 | C/T | 1 |
| SLE257 | 1 | 0 | A/G | 1 | A/A | 0 | T/G | 1 | C/T | 1 |
| SLE258 | 0 | 0 | A/G | 1 | G/A | 1 | T/G | 1 | C/C | 2 |
| SLE259 | 0 | 0 | G/G | 0 | A/A | 0 | T/G | 1 | C/T | 1 |
| SLE260 | 1 | 0 | G/G | 0 | A/A | 0 | T/T | 2 | C/C | 2 |
| SLE261 | 0 | 0 | G/G | 0 | G/A | 1 | T/G | 1 | C/T | 1 |
| SLE262 | 2 | 0 | G/G | 0 | A/A | 0 | T/T | 2 | C/C | 2 |
| SLE263 | 0 | 0 | A/A | 2 | G/A | 1 | G/G | 0 | C/T | 1 |
| SLE264 | 1 | 1 | G/G | 0 | A/A | 0 | T/T | 2 | C/C | 2 |
| SLE265 | 0 | 1 | G/G | 0 | A/A | 0 | T/T | 2 | C/C | 2 |
| SLE266 | 1 | 0 | A/G | 1 | A/A | 0 | T/G | 1 | C/T | 1 |
| SLE267 | 1 | 0 | G/G | 0 | A/A | 0 | T/G | 1 | C/T | 1 |
| SLE268 | 0 | 0 | G/G | 0 | A/A | 0 | G/G | 0 | T/T | 0 |
| SLE269 | 0 | 0 | A/A | 2 | G/A | 1 | G/G | 0 | C/T | 1 |
| SLE270 | 0 | 1 | A/G | 1 | A/A | 0 | T/G | 1 | C/T | 1 |
| SLE271 | 0 | 0 | A/G | 1 | A/A | 0 | G/G | 0 | T/T | 0 |
| SLE272 | 0 | 0 | A/G | 1 | A/A | 0 | G/G | 0 | T/T | 0 |
| SLE273 | 0 | 0 | A/G | 1 | A/A | 0 | G/G | 0 | T/T | 0 |
| SLE274 | 1 | 0 | A/G | 1 | G/A | 1 | T/G | 1 | C/C | 2 |
| SLE275 | 0 | 1 | G/G | 0 | A/A | 0 | T/G | 1 | C/T | 1 |
| SLE276 | 0 | 0 | G/G | 0 | A/A | 0 | T/G | 1 | C/T | 1 |
| SLE277 | 0 | 2 | G/G | 0 | A/A | 0 | T/T | 2 | C/C | 2 |
| SLE278 | 0 | 0 | A/A | 2 | A/A | 0 | G/G | 0 | T/T | 0 |
| SLE279 | 0 | 1 | G/G | 0 | A/A | 0 | T/G | 1 | C/T | 1 |
| SLE280 | 1 | 0 | G/G | 0 | A/A | 0 | T/G | 1 | C/T | 1 |
| SLE281 | 0 | 1 | G/G | 0 | A/A | 0 | T/T | 2 | C/C | 2 |
| SLE282 | 1 | 0 | A/G | 1 | A/A | 0 | T/G | 1 | C/T | 1 |
| SLE283 | 1 | 0 | A/G | 1 | A/A | 0 | T/G | 1 | C/T | 1 |
| SLE284 | 0 | 0 | G/G | 0 | A/A | 0 | G/G | 0 | T/T | 0 |
| SLE285 | 0 | 1 | A/G | 1 | A/A | 0 | T/G | 1 | C/T | 1 |
| SLE286 | 1 | 0 | A/G | 1 | A/A | 0 | T/G | 1 | C/T | 1 |
| SLE287 | 0 | 1 | G/G | 0 | A/A | 0 | T/G | 1 | C/T | 1 |
| SLE288 | 0 | 0 | G/G | 0 | A/A | 0 | G/G | 0 | T/T | 0 |
| SLE289 | 0 | 1 | G/G | 0 | A/A | 0 | T/T | 2 | C/C | 2 |

| | | | | | | | | | | |
|---|---|---|---|---|---|---|---|---|---|---|
| SLE290 | 0 | 0 | G/G | 0 | A/A | 0 | G/G | 0 | T/T | 0 |
| SLE291 | 0 | 0 | A/G | 1 | A/A | 0 | T/G | 1 | C/T | 1 |
| SLE292 | 0 | 0 | A/G | 1 | G/A | 1 | T/G | 1 | C/C | 2 |
| SLE293 | 1 | 0 | A/G | 1 | A/A | 0 | T/G | 1 | C/T | 1 |
| SLE294 | 1 | 0 | G/G | 0 | A/A | 0 | T/G | 1 | C/T | 1 |
| SLE295 | 0 | 0 | G/G | 0 | A/A | 0 | G/G | 0 | T/T | 0 |
| SLE296 | 0 | 0 | A/G | 1 | A/A | 0 | T/G | 1 | C/T | 1 |
| SLE297 | 0 | 2 | G/G | 0 | A/A | 0 | T/T | 2 | C/C | 2 |
| SLE298 | 0 | 1 | G/G | 0 | A/A | 0 | T/G | 1 | C/T | 1 |
| SLE299 | 0 | 0 | G/G | 0 | A/A | 0 | G/G | 0 | T/T | 0 |
| SLE300 | 0 | 0 | A/G | 1 | A/A | 0 | G/G | 0 | T/T | 0 |
| SLE301 | 0 | 1 | A/G | 1 | A/A | 0 | T/G | 1 | C/T | 1 |
| SLE302 | 0 | 0 | G/G | 0 | A/A | 0 | T/G | 1 | C/T | 1 |
| SLE303 | 0 | 0 | G/G | 0 | A/A | 0 | T/G | 1 | C/T | 1 |
| SLE304 | 0 | 0 | A/G | 1 | A/A | 0 | T/G | 1 | C/T | 1 |
| SLE305 | 1 | 0 | A/G | 1 | A/A | 0 | T/G | 1 | C/T | 1 |
| SLE306 | 0 | 0 | G/G | 0 | A/A | 0 | G/G | 0 | T/T | 0 |
| SLE307 | 1 | 0 | A/G | 1 | A/A | 0 | T/G | 1 | C/T | 1 |
| SLE308 | 0 | 1 | G/G | 0 | A/A | 0 | T/G | 1 | C/T | 1 |
| SLE309 | 0 | 1 | G/G | 0 | A/A | 0 | T/G | 1 | C/T | 1 |
| SLE310 | 0 | 0 | A/G | 1 | A/A | 0 | G/G | 0 | T/T | 0 |
| SLE311 | 1 | 0 | A/A | 2 | A/A | 0 | G/G | 0 | T/T | 0 |
| SLE312 | 0 | 0 | A/G | 1 | A/A | 0 | G/G | 0 | T/T | 0 |
| SLE313 | 0 | 0 | A/G | 1 | A/A | 0 | G/G | 0 | T/T | 0 |
| SLE314 | 0 | 1 | G/G | 0 | A/A | 0 | T/G | 1 | C/T | 1 |
| SLE315 | 0 | 0 | A/G | 1 | A/A | 0 | G/G | 0 | T/T | 0 |
| SLE316 | 0 | 2 | G/G | 0 | A/A | 0 | T/T | 2 | C/C | 2 |
| SLE317 | 0 | 0 | A/G | 1 | A/A | 0 | G/G | 0 | T/T | 0 |
| SLE318 | 0 | 0 | A/A | 2 | A/A | 0 | G/G | 0 | T/T | 0 |
| SLE319 | 1 | 0 | G/G | 0 | A/A | 0 | T/G | 1 | C/T | 1 |
| SLE320 | 1 | 0 | G/G | 0 | A/A | 0 | T/G | 1 | C/T | 1 |
| SLE321 | 1 | 0 | A/G | 1 | A/A | 0 | T/G | 1 | C/T | 1 |
| SLE322 | 1 | 1 | G/G | 0 | A/A | 0 | T/T | 2 | C/C | 2 |
| SLE323 | 0 | 0 | A/G | 1 | A/A | 0 | G/G | 0 | T/T | 0 |
| SLE324 | 1 | 1 | G/G | 0 | A/A | 0 | T/T | 2 | C/C | 2 |
| SLE325 | 0 | 0 | A/A | 2 | A/A | 0 | G/G | 0 | T/T | 0 |
| SLE326 | 0 | 0 | A/A | 2 | G/A | 1 | G/G | 0 | C/T | 1 |
| SLE327 | 1 | 0 | G/G | 0 | A/A | 0 | T/G | 1 | C/T | 1 |
| SLE328 | 0 | 0 | A/G | 1 | A/A | 0 | G/G | 0 | T/T | 0 |
| SLE329 | 0 | 0 | A/A | 2 | A/A | 0 | G/G | 0 | T/T | 0 |
| SLE330 | 0 | 1 | A/G | 1 | A/A | 0 | T/G | 1 | C/T | 1 |
| SLE331 | 1 | 0 | G/G | 0 | A/A | 0 | T/G | 1 | C/T | 1 |
| SLE332 | 2 | 0 | G/G | 0 | A/A | 0 | T/T | 2 | C/C | 2 |
| SLE333 | 0 | 0 | G/G | 0 | G/G | 2 | G/G | 0 | T/T | 0 |
| SLE334 | 0 | 0 | A/G | 1 | A/A | 0 | G/G | 0 | T/T | 0 |
| SLE335 | 0 | 1 | G/G | 0 | A/A | 0 | T/G | 1 | C/T | 1 |
| SLE336 | 0 | 1 | A/G | 1 | A/A | 0 | T/G | 1 | C/T | 1 |
| SLE337 | 0 | 0 | A/A | 2 | G/A | 1 | G/G | 0 | C/T | 1 |
| SLE338 | 0 | 0 | A/G | 1 | A/A | 0 | G/G | 0 | T/T | 0 |

| | | | | | | | | | | |
|---|---|---|---|---|---|---|---|---|---|---|
| SLE339 | 0 | 0 | A/G | 1 | A/A | 0 | G/G | 0 | T/T | 0 |
| SLE340 | 0 | 1 | G/G | 0 | A/A | 0 | T/G | 1 | C/T | 1 |
| SLE341 | 0 | 0 | G/G | 0 | A/A | 0 | G/G | 0 | T/T | 0 |
| SLE342 | 0 | 0 | A/G | 1 | A/A | 0 | G/G | 0 | T/T | 0 |
| SLE343 | 1 | 0 | G/G | 0 | A/A | 0 | T/T | 2 | C/C | 2 |
| SLE344 | 0 | 1 | G/G | 0 | A/A | 0 | T/G | 1 | C/T | 1 |
| SLE345 | 0 | 0 | A/G | 1 | A/A | 0 | G/G | 0 | T/T | 0 |
| SLE346 | 1 | 1 | G/G | 0 | A/A | 0 | T/T | 2 | C/C | 2 |
| SLE347 | 0 | 1 | G/G | 0 | A/A | 0 | T/G | 1 | C/T | 1 |
| SLE348 | 1 | 0 | G/G | 0 | A/A | 0 | T/G | 1 | C/T | 1 |
| SLE349 | 0 | 0 | A/G | 1 | A/A | 0 | G/G | 0 | T/T | 0 |
| SLE350 | 0 | 1 | G/G | 0 | A/A | 0 | T/G | 1 | C/T | 1 |
| SLE351 | 0 | 1 | A/G | 1 | A/A | 0 | T/G | 1 | C/T | 1 |
| SLE352 | 2 | 0 | G/G | 0 | A/A | 0 | T/T | 2 | C/C | 2 |
| SLE353 | 0 | 0 | G/G | 0 | A/A | 0 | G/G | 0 | T/T | 0 |
| SLE354 | 0 | 0 | G/G | 0 | G/G | 2 | G/G | 0 | T/T | 0 |
| SLE355 | 1 | 0 | A/G | 1 | A/A | 0 | T/G | 1 | C/T | 1 |
| SLE356 | 1 | 0 | G/G | 0 | A/A | 0 | T/G | 1 | C/T | 1 |
| SLE357 | 0 | 0 | A/G | 1 | A/A | 0 | G/G | 0 | T/T | 0 |
| SLE358 | 0 | 1 | G/G | 0 | A/A | 0 | T/G | 1 | C/T | 1 |
| SLE359 | 0 | 0 | G/G | 0 | A/A | 0 | T/G | 1 | C/T | 1 |
| SLE360 | 0 | 0 | A/G | 1 | G/A | 1 | G/G | 0 | C/T | 1 |
| SLE361 | 0 | 0 | A/G | 1 | A/A | 0 | G/G | 0 | T/T | 0 |
| SLE362 | 0 | 0 | A/A | 2 | A/A | 0 | G/G | 0 | T/T | 0 |
| SLE363 | 0 | 0 | A/G | 1 | A/A | 0 | G/G | 0 | T/T | 0 |
| SLE364 | 0 | 0 | A/G | 1 | A/A | 0 | G/G | 0 | T/T | 0 |
| SLE365 | 0 | 0 | A/G | 1 | A/A | 0 | G/G | 0 | T/T | 0 |
| SLE366 | 1 | 0 | A/G | 1 | G/A | 1 | T/G | 1 | C/C | 2 |
| SLE367 | 0 | 1 | A/G | 1 | A/A | 0 | T/G | 1 | C/T | 1 |
| SLE368 | 0 | 0 | G/G | 0 | A/A | 0 | T/G | 1 | C/T | 1 |
| SLE369 | 1 | 0 | A/G | 1 | A/A | 0 | T/G | 1 | C/T | 1 |
| SLE370 | 0 | 1 | A/G | 1 | A/A | 0 | T/G | 1 | C/T | 1 |
| SLE371 | 0 | 0 | A/G | 1 | G/A | 1 | G/G | 0 | C/T | 1 |
| SLE372 | 0 | 1 | A/G | 1 | A/A | 0 | T/G | 1 | C/T | 1 |
| SLE373 | 0 | 0 | A/G | 1 | A/A | 0 | G/G | 0 | T/T | 0 |
| SLE374 | 1 | 0 | G/G | 0 | A/A | 0 | T/G | 1 | C/T | 1 |
| SLE375 | 1 | 0 | A/G | 1 | A/A | 0 | T/G | 1 | C/T | 1 |
| SLE376 | 0 | 0 | A/A | 2 | A/A | 0 | G/G | 0 | T/T | 0 |
| SLE377 | 1 | 0 | A/G | 1 | A/A | 0 | T/G | 1 | C/T | 1 |
| SLE378 | 0 | 0 | A/A | 2 | G/A | 1 | G/G | 0 | C/T | 1 |
| SLE379 | 0 | 1 | G/G | 0 | A/A | 0 | T/G | 1 | C/T | 1 |
| SLE380 | 0 | 0 | G/G | 0 | A/A | 0 | G/G | 0 | T/T | 0 |
| SLE381 | 0 | 0 | A/A | 2 | A/A | 0 | G/G | 0 | T/T | 0 |
| SLE382 | 0 | 0 | A/A | 2 | A/A | 0 | G/G | 0 | T/T | 0 |
| SLE383 | 0 | 0 | G/G | 0 | A/A | 0 | G/G | 0 | T/T | 0 |
| SLE384 | 0 | 0 | A/G | 1 | A/A | 0 | G/G | 0 | T/T | 0 |
| SLE385 | 0 | 0 | G/G | 0 | A/A | 0 | T/G | 1 | C/T | 1 |
| SLE386 | 1 | 0 | A/G | 1 | A/A | 0 | T/G | 1 | C/T | 1 |
| SLE387 | 0 | 0 | A/G | 1 | G/A | 1 | G/G | 0 | C/T | 1 |

| | | | | | | | | | | |
|---|---|---|---|---|---|---|---|---|---|---|
| SLE388 | 1 | 0 | A/G | 1 | G/A | 1 | T/G | 1 | C/C | 2 |
| SLE389 | 0 | 0 | A/A | 2 | G/A | 1 | G/G | 0 | C/T | 1 |
| SLE390 | 0 | 1 | G/G | 0 | A/A | 0 | T/G | 1 | C/T | 1 |
| SLE391 | 0 | 0 | A/G | 1 | A/A | 0 | G/G | 0 | T/T | 0 |
| SLE392 | 1 | 0 | A/G | 1 | A/A | 0 | T/G | 1 | C/T | 1 |
| SLE393 | 1 | 0 | G/G | 0 | A/A | 0 | T/T | 2 | C/C | 2 |
| SLE394 | 0 | 0 | G/G | 0 | A/A | 0 | G/G | 0 | T/T | 0 |
| SLE395 | 0 | 0 | G/G | 0 | A/A | 0 | G/G | 0 | T/T | 0 |
| SLE396 | 0 | 0 | G/G | 0 | A/A | 0 | T/G | 1 | C/T | 1 |
| SLE397 | 1 | 0 | A/G | 1 | A/A | 0 | T/G | 1 | C/T | 1 |
| SLE398 | 0 | 0 | G/G | 0 | A/A | 0 | G/G | 0 | T/T | 0 |
| SLE399 | 0 | 1 | G/G | 0 | A/A | 0 | T/G | 1 | C/T | 1 |
| SLE400 | 0 | 0 | G/G | 0 | A/A | 0 | G/G | 0 | T/T | 0 |
| SLE401 | 1 | 0 | G/G | 0 | G/A | 1 | T/G | 1 | C/T | 1 |
| SLE402 | 0 | 0 | A/A | 2 | G/A | 1 | G/G | 0 | C/T | 1 |
| SLE403 | 2 | 0 | G/G | 0 | A/A | 0 | T/T | 2 | C/C | 2 |
| SLE404 | 0 | 0 | G/G | 0 | G/A | 1 | G/G | 0 | T/T | 0 |
| SLE405 | 0 | 0 | A/G | 1 | A/A | 0 | G/G | 0 | T/T | 0 |
| SLE406 | 0 | 0 | G/G | 0 | A/A | 0 | T/G | 1 | C/T | 1 |
| SLE407 | 0 | 0 | G/G | 0 | A/A | 0 | G/G | 0 | T/T | 0 |
| SLE408 | 0 | 2 | G/G | 0 | A/A | 0 | T/T | 2 | C/C | 2 |
| SLE409 | 0 | 0 | A/G | 1 | A/A | 0 | G/G | 0 | T/T | 0 |
| SLE410 | 0 | 0 | A/G | 1 | A/A | 0 | G/G | 0 | T/T | 0 |
| SLE411 | 0 | 0 | G/G | 0 | A/A | 0 | G/G | 0 | T/T | 0 |
| SLE412 | 0 | 0 | G/G | 0 | A/A | 0 | T/G | 1 | C/T | 1 |
| SLE413 | 0 | 0 | G/G | 0 | A/A | 0 | G/G | 0 | T/T | 0 |
| SLE414 | 0 | 0 | A/A | 2 | A/A | 0 | G/G | 0 | T/T | 0 |
| SLE415 | 1 | 0 | A/G | 1 | A/A | 0 | T/G | 1 | C/T | 1 |
| SLE416 | 0 | 0 | G/G | 0 | A/A | 0 | G/G | 0 | T/T | 0 |
| SLE417 | 0 | 0 | A/G | 1 | A/A | 0 | G/G | 0 | T/T | 0 |
| SLE418 | 0 | 0 | G/G | 0 | A/A | 0 | G/G | 0 | T/T | 0 |
| SLE419 | 1 | 0 | G/G | 0 | A/A | 0 | T/G | 1 | C/T | 1 |
| SLE420 | 0 | 0 | A/A | 2 | A/A | 0 | G/G | 0 | T/T | 0 |
| SLE421 | 1 | 0 | G/G | 0 | A/A | 0 | T/G | 1 | C/T | 1 |
| SLE422 | 0 | 0 | G/G | 0 | G/A | 1 | G/G | 0 | T/T | 0 |
| SLE423 | 0 | 1 | G/G | 0 | A/A | 0 | T/T | 2 | C/C | 2 |
| SLE424 | 1 | 0 | G/G | 0 | A/A | 0 | T/T | 2 | C/C | 2 |
| SLE425 | 0 | 0 | A/G | 1 | A/A | 0 | G/G | 0 | T/T | 0 |
| SLE426 | 0 | 0 | A/G | 1 | A/A | 0 | G/G | 0 | T/T | 0 |
| SLE427 | 0 | 1 | G/G | 0 | A/A | 0 | T/G | 1 | C/T | 1 |
| SLE428 | 0 | 0 | A/A | 2 | A/A | 0 | G/G | 0 | T/T | 0 |
| SLE429 | 1 | 0 | A/G | 1 | A/A | 0 | T/G | 1 | C/T | 1 |
| SLE430 | 0 | 0 | G/G | 0 | G/A | 1 | G/G | 0 | T/T | 0 |
| SLE431 | 0 | 0 | G/G | 0 | A/A | 0 | G/G | 0 | T/T | 0 |
| SLE432 | 0 | 0 | G/G | 0 | A/A | 0 | T/G | 1 | C/T | 1 |
| SLE433 | 0 | 0 | A/G | 1 | G/A | 1 | G/G | 0 | C/T | 1 |
| SLE434 | 0 | 1 | G/G | 0 | G/A | 1 | T/G | 1 | C/T | 1 |
| SLE435 | 0 | 0 | G/G | 0 | A/A | 0 | G/G | 0 | T/T | 0 |
| SLE436 | 1 | 0 | G/G | 0 | G/A | 1 | T/G | 1 | C/T | 1 |

| | | | | | | | | | | |
|---|---|---|---|---|---|---|---|---|---|---|
| SLE437 | 0 | 0 | G/G | 0 | A/A | 0 | G/G | 0 | T/T | 0 |
| SLE438 | 1 | 0 | A/G | 1 | A/A | 0 | T/G | 1 | C/T | 1 |
| SLE439 | 0 | 0 | A/G | 1 | A/A | 0 | T/G | 1 | C/T | 1 |
| SLE440 | 0 | 0 | G/G | 0 | A/A | 0 | G/G | 0 | T/T | 0 |
| SLE441 | 0 | 0 | A/G | 1 | G/A | 1 | G/G | 0 | C/T | 1 |
| SLE442 | 0 | 0 | G/G | 0 | A/A | 0 | T/G | 1 | C/T | 1 |
| HC1 | 0 | 0 | G/G | 0 | G/A | 1 | G/G | 0 | T/T | 0 |
| HC2 | 0 | 0 | A/G | 1 | G/A | 1 | G/G | 0 | T/T | 0 |
| HC3 | 0 | 0 | G/G | 0 | A/A | 0 | G/G | 0 | T/T | 0 |
| HC4 | 0 | 0 | A/G | 1 | A/A | 0 | T/G | 1 | C/T | 1 |
| HC5 | 0 | 1 | G/G | 0 | A/A | 0 | T/G | 1 | C/T | 1 |
| HC6 | 0 | 0 | A/G | 1 | A/A | 0 | G/G | 0 | T/T | 0 |
| HC7 | 0 | 0 | A/G | 1 | A/A | 0 | G/G | 0 | T/T | 0 |
| HC8 | 0 | 0 | A/G | 1 | A/A | 0 | G/G | 0 | T/T | 0 |
| HC9 | 0 | 0 | A/A | 2 | A/A | 0 | G/G | 0 | T/T | 0 |
| HC10 | 0 | 0 | G/G | 0 | A/A | 0 | T/G | 1 | C/T | 1 |
| HC11 | 0 | 0 | A/G | 1 | G/A | 1 | G/G | 0 | C/T | 1 |
| HC12 | 0 | 0 | A/A | 2 | A/A | 0 | G/G | 0 | T/T | 0 |
| HC13 | 1 | 0 | G/G | 0 | G/A | 1 | T/G | 1 | C/T | 1 |
| HC14 | 1 | 0 | A/G | 1 | A/A | 0 | T/G | 1 | C/T | 1 |
| HC15 | 0 | 0 | A/G | 1 | G/A | 1 | G/G | 0 | C/T | 1 |
| HC16 | 0 | 0 | A/G | 1 | G/A | 1 | G/G | 0 | C/T | 1 |
| HC17 | 2 | 0 | G/G | 0 | A/A | 0 | T/T | 2 | C/C | 2 |
| HC18 | 0 | 0 | A/G | 1 | A/A | 0 | G/G | 0 | T/T | 0 |
| HC19 | 0 | 1 | G/G | 0 | A/A | 0 | T/G | 1 | C/T | 1 |
| HC20 | 0 | 0 | A/A | 2 | A/A | 0 | G/G | 0 | T/T | 0 |
| HC21 | 0 | 0 | A/G | 1 | A/A | 0 | G/G | 0 | T/T | 0 |
| HC22 | 0 | 0 | A/G | 1 | A/A | 0 | T/G | 1 | C/T | 1 |
| HC23 | 0 | 0 | G/G | 0 | A/A | 0 | G/G | 0 | T/T | 0 |
| HC24 | 0 | 0 | A/G | 1 | A/A | 0 | G/G | 0 | T/T | 0 |
| HC25 | 0 | 0 | A/G | 1 | A/A | 0 | G/G | 0 | T/T | 0 |
| HC26 | 0 | 0 | A/G | 1 | A/A | 0 | G/G | 0 | T/T | 0 |
| HC27 | 0 | 0 | A/A | 2 | A/A | 0 | G/G | 0 | T/T | 0 |
| HC28 | 0 | 1 | G/G | 0 | A/A | 0 | T/G | 1 | C/T | 1 |
| HC29 | 2 | 0 | G/G | 0 | A/A | 0 | T/T | 2 | C/C | 2 |
| HC30 | 0 | 1 | G/G | 0 | A/A | 0 | T/G | 1 | C/T | 1 |
| HC31 | 0 | 0 | G/G | 0 | A/A | 0 | T/G | 1 | C/T | 1 |
| HC32 | 0 | 0 | G/G | 0 | G/A | 1 | G/G | 0 | T/T | 0 |
| HC33 | 0 | 0 | A/G | 1 | A/A | 0 | T/G | 1 | C/T | 1 |
| HC34 | 0 | 0 | G/G | 0 | G/A | 1 | T/G | 1 | C/T | 1 |
| HC35 | 0 | 0 | A/G | 1 | A/A | 0 | G/G | 0 | T/T | 0 |
| HC36 | 0 | 1 | A/G | 1 | A/A | 0 | T/G | 1 | C/T | 1 |
| HC37 | 0 | 0 | A/G | 1 | G/A | 1 | G/G | 0 | C/T | 1 |
| HC38 | 0 | 0 | G/G | 0 | A/A | 0 | G/G | 0 | T/T | 0 |
| HC39 | 0 | 1 | A/G | 1 | A/A | 0 | T/G | 1 | C/T | 1 |
| HC40 | 0 | 0 | A/G | 1 | A/A | 0 | T/G | 1 | C/T | 1 |
| HC41 | 0 | 0 | A/G | 1 | A/A | 0 | G/G | 0 | T/T | 0 |
| HC42 | 0 | 0 | G/G | 0 | G/A | 1 | T/G | 1 | C/T | 1 |
| HC43 | 0 | 0 | A/A | 2 | A/A | 0 | G/G | 0 | T/T | 0 |

| | | | | | | | | | | |
|---|---|---|---|---|---|---|---|---|---|---|
| HC44 | 1 | 0 | G/G | 0 | G/A | 1 | T/G | 1 | C/T | 1 |
| HC45 | 0 | 0 | G/G | 0 | A/A | 0 | T/G | 1 | C/T | 1 |
| HC46 | 0 | 0 | G/G | 0 | A/A | 0 | T/G | 1 | C/T | 1 |
| HC47 | 0 | 0 | A/G | 1 | A/A | 0 | G/G | 0 | T/T | 0 |
| HC48 | 0 | 0 | A/G | 1 | G/A | 1 | G/G | 0 | C/T | 1 |
| HC49 | 0 | 0 | A/G | 1 | A/A | 0 | G/G | 0 | T/T | 0 |
| HC50 | 0 | 0 | G/G | 0 | A/A | 0 | T/G | 1 | C/T | 1 |
| HC51 | 0 | 1 | G/G | 0 | A/A | 0 | T/G | 1 | C/T | 1 |
| HC52 | 0 | 1 | G/G | 0 | A/A | 0 | T/G | 1 | C/T | 1 |
| HC53 | 0 | 0 | G/G | 0 | A/A | 0 | G/G | 0 | T/T | 0 |
| HC54 | 0 | 0 | A/G | 1 | G/A | 1 | G/G | 0 | T/T | 0 |
| HC55 | 0 | 0 | G/G | 0 | A/A | 0 | G/G | 0 | T/T | 0 |
| HC56 | 0 | 1 | G/G | 0 | A/A | 0 | T/G | 1 | C/T | 1 |
| HC57 | 0 | 0 | G/G | 0 | A/A | 0 | G/G | 0 | T/T | 0 |
| HC58 | 0 | 0 | G/G | 0 | A/A | 0 | G/G | 0 | T/T | 0 |
| HC59 | 0 | 0 | G/G | 0 | G/A | 1 | G/G | 0 | T/T | 0 |
| HC60 | 0 | 0 | A/A | 2 | G/A | 1 | G/G | 0 | C/T | 1 |
| HC61 | 0 | 0 | A/G | 1 | A/A | 0 | G/G | 0 | T/T | 0 |
| HC62 | 0 | 1 | A/G | 1 | G/A | 1 | T/G | 1 | C/C | 2 |
| HC63 | 0 | 0 | G/G | 0 | G/G | 2 | G/G | 0 | T/T | 0 |
| HC64 | 0 | 0 | G/G | 0 | G/A | 1 | G/G | 0 | T/T | 0 |
| HC65 | 0 | 1 | G/G | 0 | A/A | 0 | T/G | 1 | C/T | 1 |
| HC66 | 0 | 0 | G/G | 0 | A/A | 0 | G/G | 0 | T/T | 0 |
| HC67 | 0 | 0 | A/G | 1 | G/A | 1 | G/G | 0 | C/T | 1 |
| HC68 | 0 | 0 | A/G | 1 | G/A | 1 | G/G | 0 | T/T | 0 |
| HC69 | 0 | 0 | A/G | 1 | A/A | 0 | G/G | 0 | T/T | 0 |
| HC70 | 0 | 2 | G/G | 0 | A/A | 0 | T/T | 2 | C/C | 2 |
| HC71 | 0 | 0 | A/G | 1 | A/A | 0 | T/G | 1 | C/T | 1 |
| HC72 | 0 | 1 | G/G | 0 | A/A | 0 | T/G | 1 | C/T | 1 |
| HC73 | 0 | 0 | G/G | 0 | G/A | 1 | G/G | 0 | T/T | 0 |
| HC74 | 0 | 0 | G/G | 0 | A/A | 0 | G/G | 0 | C/T | 1 |
| HC75 | 0 | 1 | G/G | 0 | A/A | 0 | T/G | 1 | C/T | 1 |
| HC76 | 1 | 0 | G/G | 0 | A/A | 0 | T/G | 1 | C/T | 1 |
| HC77 | 0 | 0 | G/G | 0 | A/A | 0 | G/G | 0 | T/T | 0 |
| HC78 | 0 | 0 | A/G | 1 | A/A | 0 | G/G | 0 | T/T | 0 |
| HC79 | 0 | 0 | A/G | 1 | A/A | 0 | G/G | 0 | T/T | 0 |
| HC80 | 0 | 1 | G/G | 0 | A/A | 0 | T/G | 1 | C/T | 1 |
| HC81 | 0 | 0 | A/A | 2 | G/A | 1 | G/G | 0 | C/T | 1 |
| HC82 | 0 | 0 | G/G | 0 | A/A | 0 | G/G | 0 | T/T | 0 |
| HC83 | 0 | 0 | G/G | 0 | G/A | 1 | G/G | 0 | T/T | 0 |
| HC84 | 0 | 0 | A/G | 1 | A/A | 0 | G/G | 0 | T/T | 0 |
| HC85 | 1 | 0 | G/G | 0 | A/A | 0 | T/G | 1 | C/T | 1 |
| HC86 | 0 | 1 | G/G | 0 | A/A | 0 | T/G | 1 | C/T | 1 |
| HC87 | 0 | 0 | A/G | 1 | G/A | 1 | G/G | 0 | C/T | 1 |
| HC88 | 0 | 1 | G/G | 0 | A/A | 0 | T/G | 1 | C/T | 1 |
| HC89 | 0 | 0 | A/G | 1 | A/A | 0 | G/G | 0 | T/T | 0 |
| HC90 | 0 | 0 | A/A | 2 | G/A | 1 | G/G | 0 | C/T | 1 |
| HC91 | 0 | 1 | A/G | 1 | G/A | 1 | T/G | 1 | C/C | 2 |
| HC92 | 1 | 0 | G/G | 0 | A/A | 0 | T/G | 1 | C/T | 1 |

| | | | | | | | | | | |
|---|---|---|---|---|---|---|---|---|---|---|
| HC93 | 0 | 0 | A/G | 1 | G/A | 1 | G/G | 0 | C/T | 1 |
| HC94 | 1 | 0 | A/G | 1 | A/A | 0 | T/G | 1 | C/T | 1 |
| HC95 | 0 | 0 | A/G | 1 | G/A | 1 | G/G | 0 | T/T | 0 |
| HC96 | 0 | 1 | A/G | 1 | A/A | 0 | T/G | 1 | C/T | 1 |
| HC97 | 0 | 0 | A/G | 1 | A/A | 0 | G/G | 0 | T/T | 0 |
| HC98 | 0 | 0 | G/G | 0 | G/A | 1 | G/G | 0 | T/T | 0 |
| HC99 | 0 | 0 | A/G | 1 | A/A | 0 | G/G | 0 | T/T | 0 |
| HC100 | 1 | 0 | A/G | 1 | G/A | 1 | T/G | 1 | C/C | 2 |
| HC101 | 0 | 1 | G/G | 0 | A/A | 0 | T/G | 1 | C/T | 1 |
| HC102 | 0 | 0 | G/G | 0 | A/A | 0 | T/G | 1 | C/T | 1 |
| HC103 | 0 | 0 | A/G | 1 | A/A | 0 | G/G | 0 | T/T | 0 |
| HC104 | 0 | 0 | A/G | 1 | G/A | 1 | G/G | 0 | C/T | 1 |
| HC105 | 0 | 0 | A/G | 1 | A/A | 0 | G/G | 0 | T/T | 0 |
| HC106 | 1 | 0 | G/G | 0 | A/A | 0 | T/G | 1 | C/T | 1 |
| HC107 | 0 | 0 | A/G | 1 | G/A | 1 | G/G | 0 | T/T | 0 |
| HC108 | 0 | 0 | A/G | 1 | A/A | 0 | G/G | 0 | T/T | 0 |
| HC109 | 0 | 0 | A/A | 2 | G/A | 1 | G/G | 0 | C/T | 1 |
| HC110 | 0 | 0 | G/G | 0 | A/A | 0 | G/G | 0 | T/T | 0 |
| HC111 | 0 | 1 | A/G | 1 | A/A | 0 | T/G | 1 | C/T | 1 |
| HC112 | 0 | 1 | G/G | 0 | A/A | 0 | T/G | 1 | C/T | 1 |
| HC113 | 0 | 0 | A/G | 1 | A/A | 0 | G/G | 0 | T/T | 0 |
| HC114 | 0 | 0 | A/G | 1 | A/A | 0 | T/G | 1 | C/T | 1 |
| HC115 | 0 | 1 | G/G | 0 | A/A | 0 | T/G | 1 | C/T | 1 |
| HC116 | 0 | 0 | A/G | 1 | G/A | 1 | G/G | 0 | C/T | 1 |
| HC117 | 0 | 0 | G/G | 0 | A/A | 0 | G/G | 0 | T/T | 0 |
| HC118 | 0 | 0 | A/A | 2 | A/A | 0 | G/G | 0 | T/T | 0 |
| HC119 | 0 | 0 | G/G | 0 | G/A | 1 | G/G | 0 | T/T | 0 |
| HC120 | 1 | 0 | A/G | 1 | A/A | 0 | T/G | 1 | C/T | 1 |
| HC121 | 0 | 0 | G/G | 0 | G/A | 1 | G/G | 0 | T/T | 0 |
| HC122 | 0 | 0 | A/G | 1 | A/A | 0 | G/G | 0 | T/T | 0 |
| HC123 | 0 | 0 | A/G | 1 | A/A | 0 | G/G | 0 | T/T | 0 |
| HC124 | 0 | 1 | A/G | 1 | A/A | 0 | T/G | 1 | C/T | 1 |
| HC125 | 0 | 0 | G/G | 0 | A/A | 0 | G/G | 0 | T/T | 0 |
| HC126 | 0 | 0 | A/G | 1 | A/A | 0 | G/G | 0 | T/T | 0 |
| HC127 | 0 | 0 | A/A | 2 | G/A | 1 | G/G | 0 | C/T | 1 |
| HC128 | 0 | 0 | A/G | 1 | A/A | 0 | G/G | 0 | T/T | 0 |
| HC129 | 0 | 1 | G/G | 0 | A/A | 0 | T/T | 2 | C/C | 2 |
| HC130 | 0 | 0 | A/A | 2 | G/A | 1 | G/G | 0 | C/T | 1 |
| HC131 | 1 | 0 | G/G | 0 | A/A | 0 | T/G | 1 | C/T | 1 |
| HC132 | 0 | 0 | A/G | 1 | G/A | 1 | G/G | 0 | T/T | 0 |
| HC133 | 0 | 0 | A/G | 1 | A/A | 0 | G/G | 0 | T/T | 0 |
| HC134 | 0 | 0 | G/G | 0 | A/A | 0 | G/G | 0 | T/T | 0 |
| HC135 | 0 | 0 | G/G | 0 | G/A | 1 | T/G | 1 | C/T | 1 |
| HC136 | 0 | 1 | G/G | 0 | A/A | 0 | T/G | 1 | C/T | 1 |
| HC137 | 0 | 1 | G/G | 0 | A/A | 0 | T/G | 1 | C/T | 1 |
| HC138 | 0 | 1 | A/G | 1 | A/A | 0 | T/G | 1 | C/T | 1 |
| HC139 | 0 | 1 | G/G | 0 | A/A | 0 | T/G | 1 | C/T | 1 |
| HC140 | 1 | 1 | G/G | 0 | A/A | 0 | T/T | 2 | C/C | 2 |
| HC141 | 0 | 1 | G/G | 0 | A/A | 0 | T/T | 2 | C/C | 2 |

| | | | | | | | | | | |
|---|---|---|---|---|---|---|---|---|---|---|
| HC142 | 0 | 0 | G/G | 0 | G/A | 1 | G/G | 0 | T/T | 0 |
| HC143 | 0 | 1 | A/G | 1 | G/A | 1 | T/G | 1 | C/C | 2 |
| HC144 | 0 | 0 | G/G | 0 | A/A | 0 | T/G | 1 | C/T | 1 |
| HC145 | 0 | 0 | G/G | 0 | G/A | 1 | G/G | 0 | T/T | 0 |
| HC146 | 0 | 0 | G/G | 0 | A/A | 0 | T/G | 1 | C/T | 1 |
| HC147 | 0 | 0 | G/G | 0 | A/A | 0 | G/G | 0 | T/T | 0 |
| HC148 | 1 | 0 | G/G | 0 | A/A | 0 | T/G | 1 | C/T | 1 |
| HC149 | 0 | 0 | G/G | 0 | G/A | 1 | G/G | 0 | T/T | 0 |
| HC150 | 0 | 1 | G/G | 0 | A/A | 0 | T/G | 1 | C/T | 1 |
| HC151 | 0 | 0 | G/G | 0 | A/A | 0 | G/G | 0 | T/T | 0 |
| HC152 | 0 | 0 | A/G | 1 | G/A | 1 | G/G | 0 | C/T | 1 |
| HC153 | 0 | 1 | G/G | 0 | A/A | 0 | T/G | 1 | C/T | 1 |
| HC154 | 1 | 0 | G/G | 0 | A/A | 0 | T/G | 1 | C/T | 1 |
| HC155 | 0 | 1 | G/G | 0 | A/A | 0 | T/G | 1 | C/T | 1 |
| HC156 | 0 | 1 | A/G | 1 | A/A | 0 | T/G | 1 | C/T | 1 |
| HC157 | 0 | 0 | A/G | 1 | A/A | 0 | T/G | 1 | C/T | 1 |
| HC158 | 0 | 0 | A/G | 1 | G/A | 1 | G/G | 0 | T/T | 0 |
| HC159 | 0 | 0 | G/G | 0 | G/A | 1 | T/G | 1 | C/T | 1 |
| HC160 | 0 | 0 | A/G | 1 | A/A | 0 | G/G | 0 | T/T | 0 |
| HC161 | 0 | 1 | A/G | 1 | G/A | 1 | T/G | 1 | C/C | 2 |
| HC162 | 0 | 0 | G/G | 0 | A/A | 0 | G/G | 0 | T/T | 0 |
| HC163 | 0 | 0 | G/G | 0 | A/A | 0 | G/G | 0 | T/T | 0 |
| HC164 | 0 | 1 | A/G | 1 | A/A | 0 | T/G | 1 | C/T | 1 |
| HC165 | 0 | 0 | A/A | 2 | A/A | 0 | G/G | 0 | T/T | 0 |
| HC166 | 0 | 0 | G/G | 0 | G/A | 1 | T/G | 1 | C/T | 1 |
| HC167 | 0 | 0 | G/G | 0 | A/A | 0 | G/G | 0 | T/T | 0 |
| HC168 | 0 | 0 | A/G | 1 | A/A | 0 | G/G | 0 | T/T | 0 |
| HC169 | 0 | 0 | A/G | 1 | A/A | 0 | G/G | 0 | T/T | 0 |
| HC170 | 0 | 1 | G/G | 0 | A/A | 0 | T/G | 1 | C/T | 1 |
| HC171 | 0 | 0 | A/G | 1 | G/A | 1 | G/G | 0 | T/T | 0 |
| HC172 | 0 | 0 | G/G | 0 | A/A | 0 | G/G | 0 | T/T | 0 |
| HC173 | 0 | 0 | A/G | 1 | A/A | 0 | G/G | 0 | T/T | 0 |
| HC174 | 0 | 1 | A/G | 1 | A/A | 0 | T/G | 1 | C/T | 1 |
| HC175 | 0 | 0 | A/G | 1 | A/A | 0 | G/G | 0 | T/T | 0 |
| HC176 | 0 | 0 | A/A | 2 | G/G | 2 | G/G | 0 | C/C | 2 |
| HC177 | 0 | 0 | A/A | 2 | A/A | 0 | G/G | 0 | T/T | 0 |
| HC178 | 0 | 1 | G/G | 0 | A/A | 0 | T/G | 1 | C/T | 1 |
| HC179 | 0 | 0 | A/G | 1 | A/A | 0 | G/G | 0 | T/T | 0 |
| HC180 | 0 | 1 | A/G | 1 | A/A | 0 | T/G | 1 | C/T | 1 |
| HC181 | 0 | 0 | A/G | 1 | G/A | 1 | G/G | 0 | C/T | 1 |
| HC182 | 0 | 0 | A/G | 1 | G/A | 1 | G/G | 0 | T/T | 0 |
| HC183 | 0 | 2 | G/G | 0 | A/A | 0 | T/T | 2 | C/C | 2 |
| HC184 | 0 | 1 | A/G | 1 | A/A | 0 | T/G | 1 | C/T | 1 |
| HC185 | 0 | 0 | A/G | 1 | A/A | 0 | G/G | 0 | T/T | 0 |
| HC186 | 1 | 0 | G/G | 0 | A/A | 0 | T/G | 1 | C/T | 1 |
| HC187 | 0 | 0 | A/G | 1 | A/A | 0 | G/G | 0 | T/T | 0 |
| HC188 | 0 | 0 | A/A | 2 | A/A | 0 | G/G | 0 | T/T | 0 |
| HC189 | 0 | 0 | A/A | 2 | G/G | 2 | G/G | 0 | C/C | 2 |
| HC190 | 0 | 0 | A/G | 1 | A/A | 0 | G/G | 0 | T/T | 0 |

| | | | | | | | | | | |
|---|---|---|---|---|---|---|---|---|---|---|
| HC191 | 0 | 1 | G/G | 0 | A/A | 0 | T/G | 1 | C/T | 1 |
| HC192 | 0 | 0 | G/G | 0 | A/A | 0 | G/G | 0 | T/T | 0 |
| HC193 | 0 | 1 | G/G | 0 | A/A | 0 | T/G | 1 | C/T | 1 |
| HC194 | 0 | 0 | G/G | 0 | A/A | 0 | T/G | 1 | C/T | 1 |
| HC195 | 0 | 1 | A/G | 1 | A/A | 0 | T/G | 1 | C/T | 1 |
| HC196 | 0 | 0 | A/G | 1 | A/A | 0 | G/G | 0 | T/T | 0 |
| HC197 | 0 | 0 | A/A | 2 | A/A | 0 | G/G | 0 | T/T | 0 |
| HC198 | 0 | 0 | G/G | 0 | A/A | 0 | G/G | 0 | T/T | 0 |
| HC199 | 0 | 0 | A/A | 2 | G/A | 1 | G/G | 0 | C/T | 1 |
| HC200 | 0 | 1 | A/G | 1 | G/A | 1 | T/G | 1 | C/C | 2 |
| HC201 | 0 | 0 | G/G | 0 | A/A | 0 | T/G | 1 | C/T | 1 |
| HC202 | 0 | 0 | G/G | 0 | A/A | 0 | G/G | 0 | T/T | 0 |
| HC203 | 0 | 0 | G/G | 0 | A/A | 0 | T/T | 2 | C/C | 2 |
| HC204 | 0 | 0 | G/G | 0 | A/A | 0 | G/G | 0 | T/T | 0 |
| HC205 | 0 | 0 | A/G | 1 | G/A | 1 | G/G | 0 | T/T | 0 |
| HC206 | 0 | 0 | G/G | 0 | G/A | 1 | T/G | 1 | C/T | 1 |
| HC207 | 0 | 0 | A/A | 2 | A/A | 0 | G/G | 0 | T/T | 0 |
| HC208 | 1 | 0 | G/G | 0 | A/A | 0 | T/G | 1 | C/T | 1 |
| HC209 | 0 | 0 | A/G | 1 | A/A | 0 | G/G | 0 | T/T | 0 |
| HC210 | 0 | 0 | G/G | 0 | A/A | 0 | G/G | 0 | T/T | 0 |
| HC211 | 0 | 0 | A/G | 1 | A/A | 0 | G/G | 0 | T/T | 0 |
| HC212 | 0 | 0 | A/G | 1 | A/A | 0 | G/G | 0 | T/T | 0 |
| HC213 | 0 | 0 | G/G | 0 | A/A | 0 | G/G | 0 | T/T | 0 |
| HC214 | 0 | 0 | A/A | 2 | A/A | 0 | G/G | 0 | T/T | 0 |
| HC215 | 2 | 0 | G/G | 0 | A/A | 0 | T/T | 2 | C/C | 2 |
| HC216 | 0 | 0 | G/G | 0 | G/A | 1 | G/G | 0 | T/T | 0 |
| HC217 | 0 | 1 | G/G | 0 | A/A | 0 | T/G | 1 | C/T | 1 |
| HC218 | 0 | 0 | G/G | 0 | A/A | 0 | G/G | 0 | T/T | 0 |
| HC219 | 0 | 1 | A/G | 1 | G/A | 1 | T/G | 1 | C/C | 2 |
| HC220 | 0 | 0 | G/G | 0 | G/A | 1 | G/G | 0 | T/T | 0 |
| HC221 | 1 | 1 | G/G | 0 | A/A | 0 | T/T | 2 | C/C | 2 |
| HC222 | 2 | 0 | G/G | 0 | A/A | 0 | T/T | 2 | C/C | 2 |
| HC223 | 0 | 0 | G/G | 0 | A/A | 0 | G/G | 0 | T/T | 0 |
| HC224 | 0 | 0 | A/G | 1 | G/A | 1 | G/G | 0 | T/T | 0 |
| HC225 | 0 | 0 | A/A | 2 | G/A | 1 | G/G | 0 | C/T | 1 |
| HC226 | 0 | 0 | A/G | 1 | A/A | 0 | G/G | 0 | T/T | 0 |
| HC227 | 0 | 0 | A/G | 1 | A/A | 0 | G/G | 0 | T/T | 0 |
| HC228 | 0 | 0 | A/A | 2 | A/A | 0 | G/G | 0 | T/T | 0 |
| HC229 | 0 | 1 | A/G | 1 | G/A | 1 | T/G | 1 | C/C | 2 |
| HC230 | 1 | 0 | G/G | 0 | A/A | 0 | T/G | 1 | C/T | 1 |
| HC231 | 0 | 1 | A/G | 1 | G/A | 1 | T/G | 1 | C/C | 2 |
| HC232 | 0 | 1 | A/G | 1 | A/A | 0 | T/G | 1 | C/T | 1 |
| HC233 | 0 | 0 | G/G | 0 | A/A | 0 | T/G | 1 | C/T | 1 |
| HC234 | 0 | 1 | A/G | 1 | A/A | 0 | T/G | 1 | C/T | 1 |
| HC235 | 0 | 0 | A/G | 1 | A/A | 0 | G/G | 0 | T/T | 0 |
| HC236 | 0 | 0 | A/G | 1 | A/A | 0 | G/G | 0 | T/T | 0 |
| HC237 | 0 | 1 | A/G | 1 | A/A | 0 | T/G | 1 | C/T | 1 |
| HC238 | 0 | 0 | A/A | 2 | A/A | 0 | G/G | 0 | T/T | 0 |
| HC239 | 0 | 0 | A/G | 1 | A/A | 0 | G/G | 0 | T/T | 0 |

| | | | | | | | | | | |
|---|---|---|---|---|---|---|---|---|---|---|
| HC240 | 0 | 1 | A/G | 1 | A/A | 0 | T/G | 1 | C/T | 1 |
| HC241 | 0 | 1 | G/G | 0 | A/A | 0 | T/G | 1 | C/T | 1 |
| HC242 | 0 | 1 | G/G | 0 | A/A | 0 | T/G | 1 | C/T | 1 |
| HC243 | 0 | 0 | A/G | 1 | G/A | 1 | G/G | 0 | C/T | 1 |
| HC244 | 0 | 0 | A/G | 1 | G/A | 1 | T/G | 1 | C/C | 2 |
| HC245 | 0 | 0 | A/A | 2 | G/A | 1 | G/G | 0 | C/T | 1 |
| HC246 | 1 | 0 | G/G | 0 | G/A | 1 | T/G | 1 | C/T | 1 |
| HC247 | 0 | 0 | A/G | 1 | A/A | 0 | G/G | 0 | T/T | 0 |
| HC248 | 0 | 0 | G/G | 0 | G/A | 1 | G/G | 0 | T/T | 0 |
| HC249 | 0 | 0 | A/G | 1 | A/A | 0 | G/G | 0 | T/T | 0 |
| HC250 | 0 | 1 | A/G | 1 | G/A | 1 | T/G | 1 | C/C | 2 |
| HC251 | 0 | 1 | G/G | 0 | A/A | 0 | T/T | 2 | C/C | 2 |
| HC252 | 0 | 0 | A/G | 1 | A/A | 0 | G/G | 0 | T/T | 0 |
| HC253 | 0 | 0 | A/G | 1 | A/A | 0 | G/G | 0 | T/T | 0 |
| HC254 | 0 | 1 | G/G | 0 | A/A | 0 | T/G | 1 | C/T | 1 |
| HC255 | 0 | 1 | A/G | 1 | A/A | 0 | T/G | 1 | C/T | 1 |
| HC256 | 0 | 0 | A/G | 1 | A/A | 0 | G/G | 0 | T/T | 0 |
| HC257 | 1 | 1 | G/G | 0 | A/A | 0 | T/T | 2 | C/C | 2 |
| HC258 | 0 | 0 | A/G | 1 | G/A | 1 | G/G | 0 | C/T | 1 |
| HC259 | 0 | 0 | A/G | 1 | A/A | 0 | G/G | 0 | T/T | 0 |
| HC260 | 1 | 0 | G/G | 0 | A/A | 0 | T/G | 1 | C/T | 1 |
| HC261 | 1 | 0 | G/G | 0 | A/A | 0 | T/G | 1 | C/T | 1 |
| HC262 | 0 | 0 | A/G | 1 | A/A | 0 | G/G | 0 | T/T | 0 |
| HC263 | 0 | 1 | A/G | 1 | G/A | 1 | T/G | 1 | C/C | 2 |
| HC264 | 0 | 0 | A/G | 1 | A/A | 0 | G/G | 0 | T/T | 0 |
| HC265 | 0 | 0 | A/G | 1 | G/A | 1 | G/G | 0 | C/T | 1 |
| HC266 | 1 | 0 | A/G | 1 | A/A | 0 | T/G | 1 | C/T | 1 |
| HC267 | 0 | 0 | A/G | 1 | A/A | 0 | G/G | 0 | T/T | 0 |
| HC268 | 0 | 0 | A/G | 1 | A/A | 0 | T/G | 1 | C/T | 1 |
| HC269 | 0 | 0 | A/G | 1 | A/A | 0 | G/G | 0 | T/T | 0 |
| HC270 | 1 | 0 | G/G | 0 | A/A | 0 | T/G | 1 | C/T | 1 |
| HC271 | 0 | 0 | G/G | 0 | A/A | 0 | G/G | 0 | T/T | 0 |
| HC272 | 0 | 1 | G/G | 0 | A/A | 0 | T/G | 1 | C/T | 1 |
| HC273 | 0 | 0 | A/A | 2 | G/A | 1 | G/G | 0 | C/T | 1 |
| HC274 | 0 | 0 | A/G | 1 | A/A | 0 | G/G | 0 | T/T | 0 |
| HC275 | 0 | 0 | G/G | 0 | A/A | 0 | G/G | 0 | T/T | 0 |
| HC276 | 0 | 0 | A/G | 1 | G/A | 1 | G/G | 0 | T/T | 0 |
| HC277 | 1 | 0 | G/G | 0 | G/A | 1 | T/G | 1 | C/T | 1 |
| HC278 | 0 | 0 | A/A | 2 | A/A | 0 | G/G | 0 | T/T | 0 |
| HC279 | 0 | 0 | G/G | 0 | A/A | 0 | G/G | 0 | T/T | 0 |
| HC280 | 0 | 0 | A/G | 1 | A/A | 0 | G/G | 0 | T/T | 0 |
| HC281 | 0 | 1 | G/G | 0 | G/A | 1 | T/G | 1 | C/T | 1 |
| HC282 | 0 | 1 | G/G | 0 | A/A | 0 | T/G | 1 | C/T | 1 |
| HC283 | 0 | 0 | A/G | 1 | G/A | 1 | G/G | 0 | C/T | 1 |
| HC284 | 0 | 0 | G/G | 0 | A/A | 0 | G/G | 0 | T/T | 0 |
| HC285 | 0 | 0 | A/G | 1 | A/A | 0 | G/G | 0 | T/T | 0 |
| HC286 | 0 | 0 | G/G | 0 | A/A | 0 | G/G | 0 | T/T | 0 |
| HC287 | 0 | 1 | A/G | 1 | G/A | 1 | T/G | 1 | C/C | 2 |
| HC288 | 0 | 1 | G/G | 0 | G/A | 1 | T/G | 1 | C/T | 1 |

| | | | | | | | | | | |
|---|---|---|---|---|---|---|---|---|---|---|
| HC289 | 0 | 0 | A/G | 1 | A/A | 0 | G/G | 0 | T/T | 0 |
| HC290 | 1 | 0 | G/G | 0 | A/A | 0 | T/G | 1 | C/T | 1 |
| HC291 | 0 | 0 | A/G | 1 | A/A | 0 | G/G | 0 | T/T | 0 |
| HC292 | 0 | 0 | A/A | 2 | G/A | 1 | G/G | 0 | C/T | 1 |
| HC293 | 0 | 0 | A/G | 1 | A/A | 0 | G/G | 0 | T/T | 0 |
| HC294 | 0 | 0 | G/G | 0 | A/A | 0 | T/G | 1 | C/T | 1 |
| HC295 | 0 | 0 | A/G | 1 | A/A | 0 | G/G | 0 | T/T | 0 |
| HC296 | 0 | 0 | A/A | 2 | G/A | 1 | G/G | 0 | C/T | 1 |
| HC297 | 0 | 0 | G/G | 0 | A/A | 0 | T/G | 1 | C/T | 1 |
| HC298 | 0 | 0 | G/G | 0 | A/A | 0 | G/G | 0 | T/T | 0 |
| HC299 | 0 | 0 | A/G | 1 | G/A | 1 | T/G | 1 | C/C | 2 |
| HC300 | 0 | 0 | A/G | 1 | A/A | 0 | G/G | 0 | T/T | 0 |
| HC301 | 0 | 1 | A/G | 1 | G/A | 1 | T/G | 1 | C/C | 2 |
| HC302 | 0 | 1 | A/G | 1 | A/A | 0 | T/G | 1 | C/T | 1 |
| HC303 | 1 | 0 | G/G | 0 | A/A | 0 | T/G | 1 | C/T | 1 |
| HC304 | 0 | 0 | A/G | 1 | A/A | 0 | G/G | 0 | T/T | 0 |
| HC305 | 0 | 0 | A/G | 1 | A/A | 0 | T/G | 1 | C/T | 1 |
| HC306 | 0 | 1 | G/G | 0 | A/A | 0 | T/G | 1 | C/T | 1 |
| HC307 | 0 | 0 | A/A | 2 | A/A | 0 | G/G | 0 | T/T | 0 |
| HC308 | 0 | 1 | A/G | 1 | G/A | 1 | T/G | 1 | C/C | 2 |
| HC309 | 0 | 0 | G/G | 0 | A/A | 0 | T/G | 1 | C/T | 1 |
| HC310 | 0 | 1 | G/G | 0 | A/A | 0 | T/G | 1 | C/T | 1 |
| HC311 | 2 | 0 | G/G | 0 | A/A | 0 | T/T | 2 | C/C | 2 |
| HC312 | 0 | 0 | A/G | 1 | A/A | 0 | G/G | 0 | T/T | 0 |
| HC313 | 1 | 0 | A/A | 2 | G/A | 1 | G/G | 0 | C/T | 1 |
| HC314 | 0 | 0 | G/G | 0 | A/A | 0 | G/G | 0 | T/T | 0 |
| HC315 | 0 | 0 | A/G | 1 | G/G | 2 | G/G | 0 | C/T | 1 |
| HC316 | 0 | 0 | A/G | 1 | A/A | 0 | G/G | 0 | T/T | 0 |
| HC317 | 0 | 0 | G/G | 0 | A/A | 0 | G/G | 0 | T/T | 0 |
| HC318 | 0 | 0 | A/A | 2 | G/A | 1 | G/G | 0 | C/T | 1 |
| HC319 | 0 | 0 | A/G | 1 | A/A | 0 | G/G | 0 | T/T | 0 |
| HC320 | 0 | 0 | G/G | 0 | A/A | 0 | G/G | 0 | T/T | 0 |
| HC321 | 0 | 0 | G/G | 0 | A/A | 0 | G/G | 0 | T/T | 0 |
| HC322 | 0 | 0 | A/G | 1 | A/A | 0 | G/G | 0 | T/T | 0 |
| HC323 | 0 | 0 | A/A | 2 | A/A | 0 | G/G | 0 | T/T | 0 |
| HC324 | 0 | 0 | G/G | 0 | G/A | 1 | G/G | 0 | T/T | 0 |
| HC325 | 0 | 0 | A/A | 2 | A/A | 0 | G/G | 0 | T/T | 0 |
| HC326 | 1 | 0 | G/G | 0 | A/A | 0 | T/G | 1 | C/T | 1 |
| HC327 | 0 | 0 | G/G | 0 | A/A | 0 | G/G | 0 | T/T | 0 |
| HC328 | 0 | 0 | A/G | 1 | A/A | 0 | G/G | 0 | T/T | 0 |
| HC329 | 1 | 0 | G/G | 0 | A/A | 0 | T/G | 1 | C/T | 1 |
| HC330 | 1 | 0 | A/G | 1 | G/A | 1 | T/G | 1 | C/C | 2 |
| HC331 | 0 | 0 | G/G | 0 | G/A | 1 | G/G | 0 | T/T | 0 |
| HC332 | 0 | 2 | G/G | 0 | A/A | 0 | T/T | 2 | C/C | 2 |
| HC333 | 0 | 0 | A/G | 1 | A/A | 0 | G/G | 0 | T/T | 0 |
| HC334 | 0 | 1 | G/G | 0 | A/A | 0 | T/G | 1 | C/T | 1 |
| HC335 | 0 | 0 | A/G | 1 | A/A | 0 | G/G | 0 | T/T | 0 |
| HC336 | 0 | 0 | A/A | 2 | G/A | 1 | G/G | 0 | C/T | 1 |
| HC337 | 0 | 0 | G/G | 0 | A/A | 0 | G/G | 0 | T/T | 0 |

| | | | | | | | | | | |
|---|---|---|---|---|---|---|---|---|---|---|
| HC338 | 1 | 0 | A/G | 1 | A/A | 0 | T/G | 1 | C/T | 1 |
| HC339 | 0 | 1 | G/G | 0 | A/A | 0 | T/G | 1 | C/T | 1 |
| HC340 | 0 | 1 | A/G | 1 | A/A | 0 | T/G | 1 | C/T | 1 |
| HC341 | 0 | 1 | G/G | 0 | A/A | 0 | T/G | 1 | C/T | 1 |
| HC342 | 0 | 1 | G/G | 0 | A/A | 0 | T/G | 1 | C/T | 1 |
| HC343 | 0 | 0 | A/G | 1 | A/A | 0 | T/G | 1 | C/T | 1 |
| HC344 | 1 | 1 | G/G | 0 | A/A | 0 | T/T | 2 | C/C | 2 |
| HC345 | 0 | 0 | A/G | 1 | G/G | 2 | G/G | 0 | C/T | 1 |
| HC346 | 0 | 0 | A/G | 1 | A/A | 0 | G/G | 0 | T/T | 0 |
| HC347 | 0 | 1 | G/G | 0 | G/A | 1 | T/G | 1 | C/T | 1 |
| HC348 | 1 | 0 | A/G | 1 | A/A | 0 | T/G | 1 | C/C | 2 |
| HC349 | 0 | 0 | G/G | 0 | A/A | 0 | T/G | 1 | C/T | 1 |
| HC350 | 0 | 0 | A/G | 1 | A/A | 0 | G/G | 0 | T/T | 0 |
| HC351 | 1 | 0 | G/G | 0 | A/A | 0 | T/G | 1 | C/T | 1 |
| HC352 | 0 | 1 | G/G | 0 | A/A | 0 | T/T | 2 | C/C | 2 |
| HC353 | 0 | 0 | G/G | 0 | A/A | 0 | G/G | 0 | T/T | 0 |
| HC354 | 0 | 0 | A/G | 1 | A/A | 0 | G/G | 0 | T/T | 0 |
| HC355 | 0 | 0 | G/G | 0 | A/A | 0 | G/G | 0 | T/T | 0 |
| HC356 | 0 | 0 | A/G | 1 | A/A | 0 | G/G | 0 | T/T | 0 |
| HC357 | 0 | 1 | G/G | 0 | A/A | 0 | T/G | 1 | C/T | 1 |
| HC358 | 0 | 0 | G/G | 0 | A/A | 0 | G/G | 0 | T/T | 0 |
| HC359 | 0 | 0 | G/G | 0 | G/A | 1 | G/G | 0 | T/T | 0 |
| HC360 | 0 | 0 | G/G | 0 | A/A | 0 | G/G | 0 | T/T | 0 |
| HC361 | 1 | 0 | G/G | 0 | A/A | 0 | T/T | 2 | C/C | 2 |
| HC362 | 0 | 0 | A/G | 1 | A/A | 0 | G/G | 0 | T/T | 0 |
| HC363 | 0 | 1 | A/G | 1 | G/A | 1 | T/G | 1 | C/C | 2 |
| HC364 | 1 | 0 | G/G | 0 | A/A | 0 | T/G | 1 | C/T | 1 |
| HC365 | 0 | 0 | A/G | 1 | A/A | 0 | G/G | 0 | T/T | 0 |
| HC366 | 0 | 0 | A/G | 1 | A/A | 0 | G/G | 0 | T/T | 0 |
| HC367 | 0 | 2 | G/G | 0 | A/A | 0 | T/T | 2 | C/C | 2 |
| HC368 | 0 | 0 | A/G | 1 | A/A | 0 | G/G | 0 | T/T | 0 |
| HC369 | 0 | 0 | A/A | 2 | G/A | 1 | G/G | 0 | C/T | 1 |
| HC370 | 0 | 0 | A/G | 1 | A/A | 0 | G/G | 0 | T/T | 0 |
| HC371 | 0 | 0 | G/G | 0 | A/A | 0 | T/G | 1 | C/T | 1 |
| HC372 | 1 | 0 | G/G | 0 | A/A | 0 | T/G | 1 | C/T | 1 |
| HC373 | 0 | 0 | A/G | 1 | A/A | 0 | G/G | 0 | T/T | 0 |
| HC374 | 0 | 0 | A/G | 1 | A/A | 0 | G/G | 0 | T/T | 0 |
| HC375 | 0 | 0 | G/G | 0 | A/A | 0 | G/G | 0 | T/T | 0 |
| HC376 | 0 | 0 | A/A | 2 | G/A | 1 | G/G | 0 | C/T | 1 |
| HC377 | 1 | 0 | G/G | 0 | A/A | 0 | T/G | 1 | C/T | 1 |
| HC378 | 0 | 0 | A/G | 1 | A/A | 0 | T/G | 1 | C/T | 1 |
| HC379 | 0 | 0 | G/G | 0 | A/A | 0 | T/G | 1 | C/T | 1 |
| HC380 | 0 | 1 | G/G | 0 | A/A | 0 | T/T | 2 | C/C | 2 |
| HC381 | 0 | 1 | A/G | 1 | A/A | 0 | T/G | 1 | C/T | 1 |
| HC382 | 1 | 0 | G/G | 0 | G/A | 1 | T/G | 1 | C/T | 1 |
| HC383 | 0 | 0 | A/G | 1 | A/A | 0 | G/G | 0 | T/T | 0 |
| HC384 | 0 | 0 | A/G | 1 | G/A | 1 | G/G | 0 | C/T | 1 |
| HC385 | 0 | 0 | A/A | 2 | A/A | 0 | G/G | 0 | T/T | 0 |
| HC386 | 0 | 0 | G/G | 0 | A/A | 0 | G/G | 0 | T/T | 0 |

| | | | | | | | | | | |
|---|---|---|---|---|---|---|---|---|---|---|
| HC387 | 1 | 1 | G/G | 0 | A/A | 0 | T/T | 2 | C/C | 2 |
| HC388 | 1 | 0 | A/G | 1 | G/A | 1 | T/G | 1 | C/C | 2 |
| HC389 | 0 | 0 | A/G | 1 | G/G | 2 | G/G | 0 | C/T | 1 |
| HC390 | 0 | 0 | A/G | 1 | A/A | 0 | T/G | 1 | C/T | 1 |
| HC391 | 0 | 0 | A/A | 2 | A/A | 0 | G/G | 0 | T/T | 0 |
| HC392 | 0 | 0 | G/G | 0 | A/A | 0 | G/G | 0 | T/T | 0 |
| HC393 | 0 | 0 | G/G | 0 | A/A | 0 | G/G | 0 | T/T | 0 |
| HC394 | 0 | 0 | A/G | 1 | A/A | 0 | T/G | 1 | C/T | 1 |
| HC395 | 0 | 0 | A/G | 1 | G/A | 1 | G/G | 0 | C/T | 1 |
| HC396 | 0 | 0 | G/G | 0 | A/A | 0 | G/G | 0 | T/T | 0 |
| HC397 | 0 | 1 | G/G | 0 | A/A | 0 | T/G | 1 | C/T | 1 |
| HC398 | 0 | 0 | G/G | 0 | G/G | 2 | G/G | 0 | T/T | 0 |
| HC399 | 0 | 0 | G/G | 0 | A/A | 0 | G/G | 0 | T/T | 0 |
| HC400 | 0 | 1 | G/G | 0 | A/A | 0 | T/G | 1 | C/T | 1 |
| HC401 | 0 | 1 | G/G | 0 | A/A | 0 | T/G | 1 | C/T | 1 |
| HC402 | 0 | 0 | G/G | 0 | A/A | 0 | G/G | 0 | T/T | 0 |
| HC403 | 0 | 0 | G/G | 0 | A/A | 0 | G/G | 0 | T/T | 0 |
| HC404 | 0 | 2 | G/G | 0 | A/A | 0 | T/T | 2 | C/C | 2 |
| HC405 | 0 | 0 | A/A | 2 | A/A | 0 | G/G | 0 | T/T | 0 |
| HC406 | 0 | 0 | A/A | 2 | A/A | 0 | G/G | 0 | T/T | 0 |
| HC407 | 0 | 0 | A/G | 1 | A/A | 0 | G/G | 0 | T/T | 0 |
| HC408 | 0 | 0 | A/G | 1 | A/A | 0 | G/G | 0 | T/T | 0 |
| HC409 | 0 | 0 | A/G | 1 | A/A | 0 | G/G | 0 | T/T | 0 |
| HC410 | 0 | 1 | G/G | 0 | A/A | 0 | T/G | 1 | C/T | 1 |
| HC411 | 0 | 0 | G/G | 0 | A/A | 0 | G/G | 0 | T/T | 0 |
| HC412 | 1 | 0 | G/G | 0 | G/A | 1 | T/G | 1 | C/T | 1 |
| HC413 | 0 | 0 | A/G | 1 | A/A | 0 | G/G | 0 | T/T | 0 |
| HC414 | 0 | 0 | A/G | 1 | A/A | 0 | T/G | 1 | C/T | 1 |
| HC415 | 0 | 1 | G/G | 0 | A/A | 0 | T/G | 1 | C/T | 1 |
| HC416 | 0 | 0 | A/G | 1 | A/A | 0 | T/G | 1 | C/T | 1 |
| HC417 | 0 | 0 | A/G | 1 | G/A | 1 | G/G | 0 | T/T | 0 |
| HC418 | 0 | 0 | A/A | 2 | G/A | 1 | G/G | 0 | C/T | 1 |
| HC419 | 0 | 0 | A/G | 1 | A/A | 0 | G/G | 0 | T/T | 0 |
| HC420 | 0 | 0 | G/G | 0 | A/A | 0 | G/G | 0 | T/T | 0 |
| HC421 | 0 | 0 | A/G | 1 | A/A | 0 | G/G | 0 | T/T | 0 |
| HC422 | 0 | 0 | G/G | 0 | A/A | 0 | G/G | 0 | T/T | 0 |
| HC423 | 0 | 0 | G/G | 0 | A/A | 0 | T/G | 1 | C/T | 1 |
| HC424 | 0 | 1 | G/G | 0 | A/A | 0 | T/G | 1 | C/T | 1 |
| HC425 | 0 | 0 | A/A | 2 | G/A | 1 | G/G | 0 | C/T | 1 |
| HC426 | 0 | 0 | A/A | 2 | G/A | 1 | G/G | 0 | C/T | 1 |
| HC427 | 0 | 0 | G/G | 0 | A/A | 0 | G/G | 0 | T/T | 0 |
| HC428 | 1 | 0 | A/G | 1 | A/A | 0 | T/G | 1 | C/T | 1 |
| HC429 | 0 | 0 | A/A | 2 | A/A | 0 | G/G | 0 | T/T | 0 |
| HC430 | 0 | 0 | A/G | 1 | A/A | 0 | T/G | 1 | C/T | 1 |
| HC431 | 0 | 0 | A/A | 2 | A/A | 0 | G/G | 0 | T/T | 0 |
| HC432 | 0 | 0 | G/G | 0 | A/A | 0 | G/G | 0 | T/T | 0 |
| HC433 | 0 | 0 | A/G | 1 | A/A | 0 | G/G | 0 | T/T | 0 |
| HC434 | 0 | 0 | A/G | 1 | A/A | 0 | T/G | 1 | C/T | 1 |
| HC435 | 0 | 0 | A/G | 1 | A/A | 0 | G/G | 0 | T/T | 0 |

| | | | | | | | | | | |
|---|---|---|---|---|---|---|---|---|---|---|
| HC436 | 0 | 1 | G/G | 0 | A/A | 0 | T/G | 1 | C/T | 1 |
| HC437 | 0 | 0 | A/A | 2 | G/A | 1 | G/G | 0 | C/T | 1 |
| HC438 | 0 | 0 | A/G | 1 | G/A | 1 | G/G | 0 | C/T | 1 |
| HC439 | 0 | 0 | G/G | 0 | A/A | 0 | G/G | 0 | T/T | 0 |
| HC440 | 0 | 0 | G/G | 0 | A/A | 0 | T/G | 1 | C/T | 1 |
| HC441 | 0 | 0 | G/G | 0 | G/A | 1 | G/G | 0 | T/T | 0 |
| HC442 | 0 | 0 | A/A | 2 | G/A | 1 | G/G | 0 | C/T | 1 |
| HC443 | 0 | 0 | A/G | 1 | A/A | 0 | G/G | 0 | T/T | 0 |
| HC444 | 0 | 0 | A/A | 2 | A/A | 0 | G/G | 0 | T/T | 0 |
| HC445 | 0 | 1 | G/G | 0 | A/A | 0 | T/G | 1 | C/T | 1 |
| HC446 | 0 | 0 | A/G | 1 | A/A | 0 | G/G | 0 | T/T | 0 |
| HC447 | 0 | 0 | G/G | 0 | G/A | 1 | G/G | 0 | T/T | 0 |
| HC448 | 0 | 1 | A/G | 1 | A/A | 0 | T/G | 1 | C/T | 1 |
| HC449 | 0 | 0 | A/G | 1 | G/A | 1 | G/G | 0 | T/T | 0 |
| HC450 | 0 | 0 | A/G | 1 | A/A | 0 | G/G | 0 | T/T | 0 |
| HC451 | 0 | 1 | A/G | 1 | A/A | 0 | T/G | 1 | C/T | 1 |
| HC452 | 0 | 0 | G/G | 0 | A/A | 0 | G/G | 0 | T/T | 0 |
| HC453 | 0 | 0 | G/G | 0 | A/A | 0 | G/G | 0 | T/T | 0 |
| HC454 | 0 | 1 | G/G | 0 | G/A | 1 | T/G | 1 | T/T | 0 |
| HC455 | 0 | 0 | A/G | 1 | A/A | 0 | G/G | 0 | T/T | 0 |
| HC456 | 0 | 0 | G/G | 0 | A/A | 0 | G/G | 0 | T/T | 0 |
| HC457 | 0 | 0 | A/A | 2 | G/A | 1 | G/G | 0 | C/T | 1 |
| HC458 | 0 | 0 | G/G | 0 | A/A | 0 | T/G | 1 | C/T | 1 |
| HC459 | 0 | 0 | A/A | 2 | A/A | 0 | G/G | 0 | T/T | 0 |
| HC460 | 0 | 0 | G/G | 0 | A/A | 0 | G/G | 0 | T/T | 0 |
| HC461 | 0 | 0 | A/G | 1 | A/A | 0 | G/G | 0 | T/T | 0 |
| HC462 | 1 | 0 | G/G | 0 | A/A | 0 | T/G | 1 | C/T | 1 |
| HC463 | 0 | 2 | G/G | 0 | A/A | 0 | T/T | 2 | C/C | 2 |
| HC464 | 0 | 0 | A/G | 1 | A/A | 0 | G/G | 0 | T/T | 0 |
| HC465 | 0 | 0 | A/G | 1 | G/A | 1 | G/G | 0 | C/T | 1 |
| HC466 | 0 | 0 | A/G | 1 | A/A | 0 | G/G | 0 | T/T | 0 |
| HC467 | 0 | 0 | A/G | 1 | G/G | 2 | G/G | 0 | C/T | 1 |
| HC468 | 0 | 0 | A/G | 1 | A/A | 0 | G/G | 0 | T/T | 0 |
| HC469 | 0 | 0 | G/G | 0 | A/A | 0 | G/G | 0 | T/T | 0 |
| HC470 | 0 | 0 | A/G | 1 | A/A | 0 | G/G | 0 | T/T | 0 |
| HC471 | 0 | 0 | A/G | 1 | A/A | 0 | G/G | 0 | T/T | 0 |
| HC472 | 0 | 0 | G/G | 0 | A/A | 0 | G/G | 0 | T/T | 0 |
| HC473 | 0 | 1 | G/G | 0 | A/A | 0 | T/G | 1 | C/T | 1 |
| HC474 | 0 | 1 | G/G | 0 | A/A | 0 | T/G | 1 | C/T | 1 |
| HC475 | 1 | 0 | A/G | 1 | A/A | 0 | T/G | 1 | C/T | 1 |
| HC476 | 0 | 0 | A/A | 2 | G/A | 1 | G/G | 0 | C/T | 1 |
| HC477 | 0 | 0 | A/G | 1 | G/A | 1 | G/G | 0 | C/T | 1 |
| HC478 | 0 | 0 | A/G | 1 | A/A | 0 | G/G | 0 | T/T | 0 |
| HC479 | 0 | 1 | A/G | 1 | G/A | 1 | T/G | 1 | C/C | 2 |
| HC480 | 0 | 0 | G/G | 0 | A/A | 0 | T/T | 2 | C/C | 2 |
| HC481 | 0 | 1 | G/G | 0 | G/A | 1 | T/G | 1 | C/T | 1 |
| HC482 | 2 | 0 | G/G | 0 | A/A | 0 | T/T | 2 | C/C | 2 |
| HC483 | 0 | 0 | A/G | 1 | G/A | 1 | G/G | 0 | T/T | 0 |
| HC484 | 0 | 0 | A/G | 1 | A/A | 0 | G/G | 0 | T/T | 0 |

| | | | | | | | | | | |
|---|---|---|---|---|---|---|---|---|---|---|
| HC485 | 0 | 0 | A/G | 1 | A/A | 0 | G/G | 0 | T/T | 0 |
| HC486 | 1 | 1 | G/G | 0 | A/A | 0 | T/T | 2 | C/C | 2 |
| HC487 | 0 | 1 | A/G | 1 | G/A | 1 | T/G | 1 | C/C | 2 |
| HC488 | 1 | 0 | A/G | 1 | A/A | 0 | T/G | 1 | C/T | 1 |
| HC489 | 0 | 1 | A/G | 1 | G/A | 1 | T/G | 1 | C/C | 2 |
| HC490 | 0 | 0 | A/G | 1 | G/G | 2 | G/G | 0 | C/T | 1 |
| HC491 | 0 | 0 | A/G | 1 | A/A | 0 | G/G | 0 | T/T | 0 |
| HC492 | 0 | 0 | A/G | 1 | A/A | 0 | G/G | 0 | T/T | 0 |
| HC493 | 1 | 0 | G/G | 0 | A/A | 0 | T/G | 1 | C/T | 1 |
| HC494 | 1 | 0 | A/G | 1 | A/A | 0 | T/G | 1 | C/T | 1 |
| HC495 | 0 | 0 | G/G | 0 | G/A | 1 | G/G | 0 | T/T | 0 |
| HC496 | 1 | 1 | G/G | 0 | A/A | 0 | T/T | 2 | C/C | 2 |
| HC497 | 0 | 0 | G/G | 0 | A/A | 0 | G/G | 0 | T/T | 0 |
| HC498 | 0 | 0 | A/G | 1 | G/A | 1 | G/G | 0 | T/T | 0 |
| HC499 | 0 | 1 | G/G | 0 | G/A | 1 | T/G | 1 | C/T | 1 |
| HC500 | 0 | 1 | G/G | 0 | A/A | 0 | T/G | 1 | C/T | 1 |
| HC501 | 0 | 1 | A/G | 1 | G/A | 1 | T/G | 1 | C/C | 2 |
| HC502 | 0 | 0 | A/G | 1 | G/A | 1 | G/G | 0 | T/T | 0 |
| HC503 | 0 | 0 | G/G | 0 | G/A | 1 | G/G | 0 | T/T | 0 |
| HC504 | 0 | 0 | A/G | 1 | A/A | 0 | G/G | 0 | T/T | 0 |
| HC505 | 0 | 0 | G/G | 0 | A/A | 0 | G/G | 0 | T/T | 0 |
| HC506 | 1 | 0 | G/G | 0 | A/A | 0 | T/G | 1 | C/T | 1 |
| HC507 | 0 | 0 | G/G | 0 | A/A | 0 | G/G | 0 | T/T | 0 |
| HC508 | 0 | 0 | A/G | 1 | G/G | 2 | G/G | 0 | C/T | 1 |
| HC509 | 1 | 0 | A/G | 1 | A/A | 0 | T/G | 1 | C/T | 1 |
| HC510 | 0 | 0 | A/A | 2 | G/G | 2 | G/G | 0 | C/C | 2 |
| HC511 | 1 | 0 | G/G | 0 | A/A | 0 | T/G | 1 | C/T | 1 |
| HC512 | 0 | 0 | A/G | 1 | G/G | 2 | G/G | 0 | C/T | 1 |
| HC513 | 0 | 1 | A/G | 1 | G/A | 1 | T/G | 1 | C/C | 2 |
| HC514 | 0 | 0 | G/G | 0 | A/A | 0 | G/G | 0 | T/T | 0 |
| HC515 | 0 | 0 | A/G | 1 | A/A | 0 | G/G | 0 | T/T | 0 |
| HC516 | 1 | 0 | G/G | 0 | A/A | 0 | T/G | 1 | C/T | 1 |
| HC517 | 1 | 1 | G/G | 0 | A/A | 0 | T/T | 2 | C/C | 2 |
| HC518 | 0 | 1 | A/G | 1 | A/A | 0 | T/G | 1 | C/T | 1 |
| HC519 | 0 | 0 | A/G | 1 | A/A | 0 | G/G | 0 | T/T | 0 |
| HC520 | 0 | 0 | A/G | 1 | G/A | 1 | G/G | 0 | C/T | 1 |
| HC521 | 0 | 0 | A/G | 1 | A/A | 0 | G/G | 0 | T/T | 0 |
| HC522 | 0 | 0 | A/G | 1 | A/A | 0 | G/G | 0 | T/T | 0 |
| HC523 | 0 | 0 | A/A | 2 | G/A | 1 | G/G | 0 | C/T | 1 |
| HC524 | 0 | 0 | A/A | 2 | A/A | 0 | G/G | 0 | T/T | 0 |
| HC525 | 0 | 0 | A/A | 2 | A/A | 0 | G/G | 0 | T/T | 0 |
| HC526 | 0 | 0 | A/G | 1 | A/A | 0 | G/G | 0 | T/T | 0 |
| HC527 | 0 | 0 | A/G | 1 | A/A | 0 | G/G | 0 | T/T | 0 |
| HC528 | 0 | 0 | G/G | 0 | A/A | 0 | T/G | 1 | C/T | 1 |
| HC529 | 0 | 1 | G/G | 0 | A/A | 0 | T/G | 1 | C/T | 1 |
| HC530 | 0 | 2 | G/G | 0 | A/A | 0 | T/T | 2 | C/C | 2 |
| HC531 | 0 | 0 | G/G | 0 | A/A | 0 | G/G | 0 | T/T | 0 |
| HC532 | 0 | 1 | G/G | 0 | A/A | 0 | T/G | 1 | C/T | 1 |
| HC533 | 0 | 0 | G/G | 0 | A/A | 0 | G/G | 0 | T/T | 0 |

| | | | | | | | | | | |
|---|---|---|---|---|---|---|---|---|---|---|
| HC534 | 0 | 1 | G/G | 0 | A/A | 0 | T/G | 1 | C/T | 1 |
| HC535 | 0 | 0 | G/G | 0 | A/A | 0 | T/G | 1 | C/T | 1 |
| HC536 | 0 | 0 | G/G | 0 | A/A | 0 | T/G | 1 | C/T | 1 |
| HC537 | 0 | 1 | G/G | 0 | A/A | 0 | T/G | 1 | C/T | 1 |
| HC538 | 0 | 0 | A/G | 1 | A/A | 0 | G/G | 0 | T/T | 0 |
| HC539 | 0 | 0 | A/G | 1 | A/A | 0 | G/G | 0 | T/T | 0 |
| HC540 | 0 | 1 | G/G | 0 | A/A | 0 | T/G | 1 | C/T | 1 |
| HC541 | 0 | 1 | G/G | 0 | A/A | 0 | T/G | 1 | C/T | 1 |
| HC542 | 0 | 1 | G/G | 0 | A/A | 0 | T/G | 1 | C/T | 1 |
| HC543 | 0 | 0 | G/G | 0 | G/A | 1 | G/G | 0 | T/T | 0 |
| HC544 | 1 | 0 | A/G | 1 | A/A | 0 | T/G | 1 | C/T | 1 |
| HC545 | 0 | 0 | A/G | 1 | A/A | 0 | G/G | 0 | T/T | 0 |
| HC546 | 0 | 0 | A/A | 2 | G/A | 1 | G/G | 0 | C/T | 1 |
| HC547 | 0 | 1 | A/G | 1 | A/A | 0 | T/G | 1 | C/T | 1 |
| HC548 | 0 | 1 | A/G | 1 | A/A | 0 | T/G | 1 | C/T | 1 |
| HC549 | 0 | 0 | A/A | 2 | A/A | 0 | G/G | 0 | T/T | 0 |
| HC550 | 0 | 1 | G/G | 0 | A/A | 0 | T/T | 2 | C/C | 2 |
| HC551 | 0 | 0 | G/G | 0 | A/A | 0 | G/G | 0 | T/T | 0 |
| HC552 | 0 | 0 | G/G | 0 | A/A | 0 | T/G | 1 | C/T | 1 |
| HC553 | 0 | 0 | G/G | 0 | A/A | 0 | T/G | 1 | C/T | 1 |
| HC554 | 0 | 0 | A/A | 2 | A/A | 0 | G/G | 0 | T/T | 0 |
| HC555 | 0 | 1 | G/G | 0 | A/A | 0 | T/T | 2 | C/C | 2 |
| HC556 | 0 | 0 | A/G | 1 | A/A | 0 | G/G | 0 | T/T | 0 |
| HC557 | 0 | 2 | G/G | 0 | A/A | 0 | T/T | 2 | C/C | 2 |
| HC558 | 0 | 0 | A/G | 1 | A/A | 0 | G/G | 0 | T/T | 0 |
| HC559 | 0 | 0 | A/G | 1 | A/A | 0 | G/G | 0 | T/T | 0 |
| HC560 | 1 | 0 | G/G | 0 | A/A | 0 | T/G | 1 | C/T | 1 |
| HC561 | 0 | 0 | A/A | 2 | G/A | 1 | G/G | 0 | C/T | 1 |
| HC562 | 0 | 0 | A/G | 1 | G/A | 1 | G/G | 0 | T/T | 0 |
| HC563 | 0 | 0 | A/G | 1 | A/A | 0 | G/G | 0 | T/T | 0 |
| HC564 | 0 | 0 | A/G | 1 | G/A | 1 | G/G | 0 | C/T | 1 |
| HC565 | 0 | 0 | G/G | 0 | A/A | 0 | G/G | 0 | T/T | 0 |
| HC566 | 1 | 0 | A/G | 1 | A/A | 0 | T/G | 1 | C/T | 1 |
| HC567 | 0 | 0 | G/G | 0 | G/A | 1 | T/G | 1 | C/T | 1 |
| HC568 | 0 | 1 | A/G | 1 | A/A | 0 | T/G | 1 | C/T | 1 |
| HC569 | 0 | 0 | G/G | 0 | A/A | 0 | G/G | 0 | T/T | 0 |
| HC570 | 0 | 0 | A/A | 2 | A/A | 0 | G/G | 0 | T/T | 0 |
| HC571 | 0 | 0 | G/G | 0 | A/A | 0 | T/G | 1 | C/T | 1 |
| HC572 | 2 | 0 | G/G | 0 | A/A | 0 | T/T | 2 | C/C | 2 |
| HC573 | 1 | 0 | A/G | 1 | G/A | 1 | T/G | 1 | C/C | 2 |
| HC574 | 0 | 0 | G/G | 0 | A/A | 0 | G/G | 0 | T/T | 0 |
| HC575 | 0 | 0 | A/G | 1 | A/A | 0 | G/G | 0 | T/T | 0 |
| HC576 | 0 | 0 | A/G | 1 | G/A | 1 | G/G | 0 | C/T | 1 |
| HC577 | 0 | 0 | A/A | 2 | A/A | 0 | G/G | 0 | T/T | 0 |
| HC578 | 0 | 0 | G/G | 0 | A/A | 0 | G/G | 0 | T/T | 0 |
| HC579 | 0 | 0 | A/G | 1 | G/A | 1 | G/G | 0 | C/T | 1 |
| HC580 | 0 | 1 | G/G | 0 | A/A | 0 | T/G | 1 | C/T | 1 |
| HC581 | 0 | 1 | G/G | 0 | G/A | 1 | T/G | 1 | C/T | 1 |
| HC582 | 0 | 2 | G/G | 0 | A/A | 0 | T/T | 2 | C/C | 2 |

| | | | | | | | | | | |
|---|---|---|---|---|---|---|---|---|---|---|
| HC583 | 0 | 1 | A/G | 1 | A/A | 0 | T/G | 1 | C/T | 1 |
| HC584 | 0 | 0 | A/G | 1 | G/A | 1 | T/G | 1 | C/C | 2 |
| HC585 | 0 | 0 | G/G | 0 | G/A | 1 | G/G | 0 | T/T | 0 |
| HC586 | 0 | 0 | A/A | 2 | G/G | 2 | G/G | 0 | C/C | 2 |
| HC587 | 0 | 0 | A/G | 1 | A/A | 0 | G/G | 0 | T/T | 0 |
| HC588 | 0 | 0 | G/G | 0 | A/A | 0 | G/G | 0 | T/T | 0 |
| HC589 | 0 | 1 | A/G | 1 | A/A | 0 | T/G | 1 | C/T | 1 |
| HC590 | 0 | 0 | A/G | 1 | A/A | 0 | G/G | 0 | T/T | 0 |
| HC591 | 0 | 0 | A/G | 1 | A/A | 0 | G/G | 0 | T/T | 0 |
| HC592 | 1 | 0 | G/G | 0 | A/A | 0 | T/G | 1 | C/T | 1 |
| HC593 | 0 | 1 | G/G | 0 | G/A | 1 | T/G | 1 | C/T | 1 |
| HC594 | 0 | 0 | G/G | 0 | A/A | 0 | G/G | 0 | T/T | 0 |
| HC595 | 0 | 1 | A/G | 1 | A/A | 0 | T/G | 1 | C/T | 1 |
| HC596 | 0 | 0 | G/G | 0 | G/A | 1 | G/G | 0 | T/T | 0 |
| HC597 | 0 | 0 | A/G | 1 | A/A | 0 | G/G | 0 | T/T | 0 |
| HC598 | 0 | 0 | G/G | 0 | A/A | 0 | G/G | 0 | T/T | 0 |
| HC599 | 0 | 1 | A/G | 1 | A/A | 0 | T/G | 1 | C/T | 1 |
| HC600 | 0 | 0 | G/G | 0 | A/A | 0 | G/G | 0 | T/T | 0 |
| HC601 | 0 | 0 | A/G | 1 | G/A | 1 | G/G | 0 | T/T | 0 |
| HC602 | 0 | 0 | A/G | 1 | A/A | 0 | T/G | 1 | C/T | 1 |
| HC603 | 0 | 0 | G/G | 0 | A/A | 0 | G/G | 0 | T/T | 0 |
| HC604 | 0 | 0 | A/G | 1 | A/A | 0 | G/G | 0 | T/T | 0 |
| HC605 | 0 | 0 | A/G | 1 | A/A | 0 | G/G | 0 | T/T | 0 |
| HC606 | 0 | 0 | A/G | 1 | A/A | 0 | G/G | 0 | T/T | 0 |
| HC607 | 0 | 0 | A/G | 1 | A/A | 0 | G/G | 0 | T/T | 0 |
| HC608 | 0 | 0 | G/G | 0 | A/A | 0 | T/G | 1 | C/T | 1 |
| HC609 | 0 | 2 | G/G | 0 | A/A | 0 | T/T | 2 | C/C | 2 |
| HC610 | 0 | 0 | G/G | 0 | G/A | 1 | G/G | 0 | T/T | 0 |
| HC611 | 1 | 0 | G/G | 0 | A/A | 0 | T/T | 2 | C/C | 2 |
| HC612 | 1 | 0 | G/G | 0 | A/A | 0 | T/G | 1 | C/T | 1 |
| HC613 | 0 | 0 | A/G | 1 | A/A | 0 | G/G | 0 | T/T | 0 |
| HC614 | 0 | 0 | A/G | 1 | G/A | 1 | G/G | 0 | C/T | 1 |
| HC615 | 0 | 0 | G/G | 0 | G/A | 1 | G/G | 0 | T/T | 0 |
| HC616 | 0 | 0 | A/G | 1 | A/A | 0 | G/G | 0 | T/T | 0 |
| HC617 | 0 | 0 | A/G | 1 | G/A | 1 | G/G | 0 | C/T | 1 |
| HC618 | 0 | 0 | A/A | 2 | A/A | 0 | G/G | 0 | T/T | 0 |
| HC619 | 0 | 1 | A/G | 1 | A/A | 0 | T/G | 1 | C/T | 1 |
| HC620 | 0 | 1 | G/G | 0 | A/A | 0 | T/G | 1 | C/T | 1 |
| HC621 | 0 | 1 | G/G | 0 | A/A | 0 | T/G | 1 | C/T | 1 |
| HC622 | 0 | 0 | A/G | 1 | A/A | 0 | G/G | 0 | T/T | 0 |
| HC623 | 0 | 0 | G/G | 0 | G/A | 1 | G/G | 0 | T/T | 0 |
| HC624 | 0 | 0 | A/A | 2 | G/A | 1 | G/G | 0 | C/T | 1 |
| HC625 | 0 | 0 | A/G | 1 | A/A | 0 | G/G | 0 | T/T | 0 |
| HC626 | 0 | 0 | A/G | 1 | A/A | 0 | G/G | 0 | T/T | 0 |
| HC627 | 0 | 1 | G/G | 0 | A/A | 0 | T/G | 1 | C/T | 1 |
| HC628 | 0 | 1 | G/G | 0 | A/A | 0 | T/G | 1 | C/T | 1 |
| HC629 | 0 | 1 | G/G | 0 | A/A | 0 | T/T | 2 | C/C | 2 |
| HC630 | 0 | 0 | A/G | 1 | A/A | 0 | G/G | 0 | T/T | 0 |
| HC631 | 0 | 0 | A/G | 1 | A/A | 0 | G/G | 0 | T/T | 0 |

| | | | | | | | | | | |
|---|---|---|---|---|---|---|---|---|---|---|
| HC632 | 0 | 1 | A/G | 1 | G/A | 1 | T/G | 1 | C/C | 2 |
| HC633 | 0 | 0 | A/A | 2 | G/A | 1 | G/G | 0 | C/T | 1 |
| HC634 | 0 | 0 | A/G | 1 | A/A | 0 | G/G | 0 | T/T | 0 |
| HC635 | 0 | 0 | A/A | 2 | G/G | 2 | G/G | 0 | C/C | 2 |
| HC636 | 0 | 0 | A/A | 2 | G/A | 1 | G/G | 0 | C/T | 1 |
| HC637 | 0 | 0 | G/G | 0 | A/A | 0 | T/G | 1 | C/T | 1 |
| HC638 | 0 | 0 | G/G | 0 | G/A | 1 | T/G | 1 | C/T | 1 |
| HC639 | 0 | 0 | A/G | 1 | A/A | 0 | G/G | 0 | T/T | 0 |
| HC640 | 0 | 0 | A/G | 1 | G/G | 2 | G/G | 0 | C/T | 1 |
| HC641 | 0 | 0 | G/G | 0 | G/A | 1 | G/G | 0 | T/T | 0 |
| HC642 | 0 | 0 | A/G | 1 | A/A | 0 | G/G | 0 | T/T | 0 |
| HC643 | 0 | 1 | G/G | 0 | A/A | 0 | T/G | 1 | C/T | 1 |
| HC644 | 1 | 0 | A/G | 1 | A/A | 0 | T/G | 1 | C/T | 1 |
| HC645 | 0 | 0 | G/G | 0 | A/A | 0 | G/G | 0 | T/T | 0 |
| HC646 | 1 | 0 | G/G | 0 | A/A | 0 | T/T | 2 | C/C | 2 |
| HC647 | 0 | 0 | A/G | 1 | A/A | 0 | G/G | 0 | T/T | 0 |
| HC648 | 1 | 0 | A/G | 1 | A/A | 0 | T/G | 1 | C/T | 1 |
| HC649 | 0 | 0 | A/G | 1 | A/A | 0 | T/G | 1 | C/T | 1 |
| HC650 | 0 | 0 | A/G | 1 | G/A | 1 | G/G | 0 | C/T | 1 |
| HC651 | 0 | 0 | G/G | 0 | G/A | 1 | T/G | 1 | C/T | 1 |
| HC652 | 0 | 0 | A/A | 2 | G/A | 1 | G/G | 0 | C/T | 1 |
| HC653 | 0 | 0 | A/A | 2 | G/A | 1 | G/G | 0 | C/T | 1 |
| HC654 | 0 | 0 | G/G | 0 | A/A | 0 | G/G | 0 | T/T | 0 |
| HC655 | 0 | 1 | A/G | 1 | A/A | 0 | T/G | 1 | C/T | 1 |
| HC656 | 0 | 0 | A/G | 1 | A/A | 0 | G/G | 0 | T/T | 0 |
| HC657 | 0 | 0 | A/G | 1 | A/A | 0 | G/G | 0 | T/T | 0 |
| HC658 | 1 | 0 | G/G | 0 | A/A | 0 | T/G | 1 | C/T | 1 |
| HC659 | 0 | 1 | A/G | 1 | G/A | 1 | T/G | 1 | C/C | 2 |
| HC660 | 0 | 0 | G/G | 0 | A/A | 0 | G/G | 0 | T/T | 0 |
| HC661 | 0 | 0 | A/G | 1 | A/A | 0 | T/G | 1 | C/T | 1 |
| HC662 | 0 | 1 | G/G | 0 | G/A | 1 | T/G | 1 | C/T | 1 |
| HC663 | 0 | 1 | A/G | 1 | A/A | 0 | T/G | 1 | C/T | 1 |
| HC664 | 0 | 1 | G/G | 0 | A/A | 0 | T/G | 1 | C/T | 1 |
| HC665 | 0 | 1 | G/G | 0 | A/A | 0 | T/G | 1 | C/T | 1 |
| HC666 | 0 | 0 | A/A | 2 | A/A | 0 | G/G | 0 | T/T | 0 |
| HC667 | 0 | 1 | G/G | 0 | A/A | 0 | T/G | 1 | C/T | 1 |
| HC668 | 1 | 0 | G/G | 0 | A/A | 0 | T/G | 1 | C/T | 1 |
| HC669 | 0 | 0 | A/G | 1 | A/A | 0 | G/G | 0 | T/T | 0 |
| HC670 | 0 | 1 | A/G | 1 | A/A | 0 | T/G | 1 | C/T | 1 |
| HC671 | 0 | 0 | G/G | 0 | A/A | 0 | G/G | 0 | T/T | 0 |
| HC672 | 0 | 0 | A/G | 1 | A/A | 0 | T/G | 1 | C/T | 1 |
| HC673 | 0 | 0 | A/G | 1 | A/A | 0 | G/G | 0 | T/T | 0 |
| HC674 | 0 | 0 | G/G | 0 | G/A | 1 | G/G | 0 | T/T | 0 |
| HC675 | 0 | 0 | A/A | 2 | G/A | 1 | G/G | 0 | C/T | 1 |
| HC676 | 0 | 0 | A/G | 1 | A/A | 0 | G/G | 0 | T/T | 0 |
| HC677 | 1 | 0 | G/G | 0 | A/A | 0 | T/G | 1 | C/T | 1 |
| HC678 | 0 | 0 | A/G | 1 | A/A | 0 | G/G | 0 | T/T | 0 |
| HC679 | 1 | 0 | G/G | 0 | G/A | 1 | T/G | 1 | C/T | 1 |
| HC680 | 0 | 1 | G/G | 0 | A/A | 0 | T/G | 1 | C/T | 1 |

| | | | | | | | | | | |
|---|---|---|---|---|---|---|---|---|---|---|
| HC681 | 0 | 0 | G/G | 0 | G/A | 1 | G/G | 0 | T/T | 0 |
| HC682 | 0 | 0 | A/G | 1 | A/A | 0 | G/G | 0 | T/T | 0 |
| HC683 | 0 | 1 | G/G | 0 | A/A | 0 | T/G | 1 | C/T | 1 |
| HC684 | 0 | 1 | A/G | 1 | A/A | 0 | T/G | 1 | C/T | 1 |
| HC685 | 0 | 0 | G/G | 0 | G/A | 1 | G/G | 0 | T/T | 0 |
| HC686 | 0 | 1 | G/G | 0 | A/A | 0 | T/G | 1 | C/T | 1 |
| HC687 | 0 | 0 | A/G | 1 | A/A | 0 | G/G | 0 | T/T | 0 |
| HC688 | 0 | 0 | G/G | 0 | G/A | 1 | G/G | 0 | T/T | 0 |
| HC689 | 0 | 1 | A/G | 1 | A/A | 0 | T/G | 1 | C/T | 1 |
| HC690 | 0 | 0 | A/G | 1 | A/A | 0 | G/G | 0 | T/T | 0 |
| HC691 | 0 | 0 | G/G | 0 | A/A | 0 | G/G | 0 | T/T | 0 |
| HC692 | 0 | 0 | G/G | 0 | A/A | 0 | T/G | 1 | C/T | 1 |
| HC693 | 0 | 0 | G/G | 0 | A/A | 0 | G/G | 0 | T/T | 0 |
| HC694 | 1 | 0 | A/G | 1 | A/A | 0 | T/G | 1 | C/T | 1 |
| HC695 | 1 | 0 | A/G | 1 | A/A | 0 | T/G | 1 | C/T | 1 |
| HC696 | 0 | 1 | A/G | 1 | A/A | 0 | T/G | 1 | C/T | 1 |
| HC697 | 0 | 0 | A/G | 1 | G/A | 1 | G/G | 0 | C/T | 1 |
| HC698 | 0 | 0 | A/G | 1 | G/A | 1 | G/G | 0 | C/T | 1 |
| HC699 | 0 | 0 | A/G | 1 | A/A | 0 | G/G | 0 | T/T | 0 |
| HC700 | 1 | 0 | A/G | 1 | A/A | 0 | T/G | 1 | C/T | 1 |
| HC701 | 0 | 1 | A/G | 1 | A/A | 0 | T/G | 1 | C/T | 1 |
| HC702 | 0 | 0 | G/G | 0 | A/A | 0 | G/G | 0 | T/T | 0 |
| HC703 | 1 | 0 | G/G | 0 | A/A | 0 | T/T | 2 | C/C | 2 |
| HC704 | 0 | 1 | A/G | 1 | G/A | 1 | T/G | 1 | C/C | 2 |
| HC705 | 0 | 1 | G/G | 0 | G/A | 1 | T/G | 1 | C/T | 1 |
| HC706 | 0 | 0 | A/G | 1 | A/A | 0 | G/G | 0 | T/T | 0 |
| HC707 | 0 | 2 | G/G | 0 | A/A | 0 | T/T | 2 | C/C | 2 |
| HC708 | 0 | 1 | G/G | 0 | A/A | 0 | T/G | 1 | C/T | 1 |
| HC709 | 0 | 0 | G/G | 0 | A/A | 0 | G/G | 0 | T/T | 0 |
| HC710 | 0 | 0 | G/G | 0 | A/A | 0 | G/G | 0 | T/T | 0 |
| HC711 | 0 | 0 | A/G | 1 | G/A | 1 | G/G | 0 | C/T | 1 |
| HC712 | 0 | 0 | G/G | 0 | G/A | 1 | G/G | 0 | T/T | 0 |
| HC713 | 0 | 0 | A/G | 1 | A/A | 0 | G/G | 0 | T/T | 0 |
| HC714 | 0 | 0 | G/G | 0 | A/A | 0 | T/G | 1 | C/T | 1 |
| HC715 | 0 | 0 | A/A | 2 | A/A | 0 | G/G | 0 | T/T | 0 |
| HC716 | 0 | 0 | A/G | 1 | A/A | 0 | G/G | 0 | T/T | 0 |
| HC717 | 0 | 0 | G/G | 0 | A/A | 0 | G/G | 0 | T/T | 0 |
| HC718 | 1 | 0 | G/G | 0 | G/A | 1 | T/G | 1 | C/T | 1 |
| HC719 | 1 | 0 | A/G | 1 | A/A | 0 | T/G | 1 | C/T | 1 |
| HC720 | 0 | 0 | A/G | 1 | A/A | 0 | G/G | 0 | T/T | 0 |
| HC721 | 0 | 0 | A/A | 2 | G/A | 1 | G/G | 0 | C/T | 1 |
| HC722 | 1 | 0 | A/G | 1 | A/A | 0 | T/G | 1 | C/T | 1 |
| HC723 | 0 | 0 | G/G | 0 | A/A | 0 | G/G | 0 | T/T | 0 |
| HC724 | 1 | 0 | G/G | 0 | A/A | 0 | T/G | 1 | C/T | 1 |
| HC725 | 0 | 1 | A/G | 1 | G/A | 1 | T/G | 1 | C/C | 2 |
| HC726 | 0 | 1 | G/G | 0 | A/A | 0 | T/T | 2 | C/C | 2 |
| HC727 | 0 | 0 | G/G | 0 | G/A | 1 | T/G | 1 | C/T | 1 |
| HC728 | 1 | 0 | G/G | 0 | A/A | 0 | T/G | 1 | C/T | 1 |
| HC729 | 0 | 1 | G/G | 0 | A/A | 0 | T/G | 1 | C/T | 1 |

| | | | | | | | | | | |
|---|---|---|---|---|---|---|---|---|---|---|
| HC730 | 0 | 0 | A/G | 1 | A/A | 0 | G/G | 0 | T/T | 0 |
| HC731 | 0 | 1 | G/G | 0 | A/A | 0 | T/G | 1 | C/T | 1 |
| HC732 | 0 | 1 | G/G | 0 | A/A | 0 | T/T | 2 | C/C | 2 |
| HC733 | 0 | 0 | G/G | 0 | A/A | 0 | G/G | 0 | T/T | 0 |
| HC734 | 1 | 0 | G/G | 0 | A/A | 0 | T/G | 1 | C/T | 1 |
| HC735 | 0 | 1 | A/G | 1 | A/A | 0 | T/G | 1 | C/T | 1 |
| HC736 | 0 | 0 | G/G | 0 | A/A | 0 | G/G | 0 | T/T | 0 |
| HC737 | 0 | 0 | G/G | 0 | A/A | 0 | G/G | 0 | T/T | 0 |
| HC738 | 0 | 0 | G/G | 0 | A/A | 0 | T/G | 1 | C/T | 1 |
| HC739 | 1 | 0 | A/G | 1 | A/A | 0 | T/G | 1 | C/T | 1 |
| HC740 | 0 | 0 | G/G | 0 | G/A | 1 | G/G | 0 | T/T | 0 |
| HC741 | 0 | 1 | G/G | 0 | A/A | 0 | T/G | 1 | C/T | 1 |
| HC742 | 0 | 0 | G/G | 0 | A/A | 0 | G/G | 0 | T/T | 0 |
| HC743 | 0 | 0 | G/G | 0 | G/A | 1 | G/G | 0 | T/T | 0 |
| HC744 | 0 | 1 | G/G | 0 | A/A | 0 | T/G | 1 | C/T | 1 |
| HC745 | 0 | 0 | G/G | 0 | A/A | 0 | G/G | 0 | T/T | 0 |
| HC746 | 0 | 0 | A/G | 1 | A/A | 0 | T/G | 1 | C/T | 1 |
| HC747 | 1 | 0 | A/G | 1 | A/A | 0 | T/G | 1 | C/T | 1 |
| HC748 | 0 | 0 | A/G | 1 | G/A | 1 | G/G | 0 | C/T | 1 |
| HC749 | 0 | 0 | G/G | 0 | A/A | 0 | G/G | 0 | T/T | 0 |
| HC750 | 1 | 1 | G/G | 0 | A/A | 0 | T/T | 2 | C/C | 2 |
| HC751 | 0 | 0 | A/G | 1 | G/A | 1 | G/G | 0 | C/T | 1 |
| HC752 | 0 | 0 | A/G | 1 | A/A | 0 | T/G | 1 | C/T | 1 |
| HC753 | 0 | 0 | A/A | 2 | A/A | 0 | G/G | 0 | T/T | 0 |
| HC754 | 0 | 0 | A/A | 2 | G/G | 2 | G/G | 0 | C/C | 2 |
| HC755 | 0 | 1 | G/G | 0 | A/A | 0 | T/G | 1 | C/T | 1 |
| HC756 | 0 | 0 | A/G | 1 | G/A | 1 | G/G | 0 | C/T | 1 |
| HC757 | 1 | 0 | G/G | 0 | A/A | 0 | T/G | 1 | C/T | 1 |
| HC758 | 0 | 1 | G/G | 0 | A/A | 0 | T/T | 2 | C/C | 2 |
| HC759 | 0 | 1 | A/G | 1 | A/A | 0 | T/G | 1 | C/T | 1 |
| HC760 | 1 | 0 | G/G | 0 | A/A | 0 | T/G | 1 | C/T | 1 |
| HC761 | 0 | 0 | A/G | 1 | G/A | 1 | G/G | 0 | T/T | 0 |
| HC762 | 1 | 0 | A/G | 1 | A/A | 0 | T/G | 1 | C/T | 1 |
| HC763 | 1 | 0 | G/G | 0 | A/A | 0 | T/G | 1 | C/T | 1 |
| HC764 | 0 | 0 | A/A | 2 | A/A | 0 | G/G | 0 | T/T | 0 |
| HC765 | 1 | 0 | A/G | 1 | A/A | 0 | T/G | 1 | C/T | 1 |
| HC766 | 0 | 0 | A/G | 1 | A/A | 0 | G/G | 0 | T/T | 0 |
| HC767 | 0 | 1 | G/G | 0 | G/A | 1 | T/G | 1 | C/T | 1 |
| HC768 | 1 | 0 | G/G | 0 | A/A | 0 | T/T | 2 | C/C | 2 |
| HC769 | 0 | 0 | A/G | 1 | G/A | 1 | G/G | 0 | C/T | 1 |
| HC770 | 0 | 1 | G/G | 0 | A/A | 0 | T/T | 2 | C/C | 2 |
| HC771 | 0 | 0 | A/G | 1 | A/A | 0 | G/G | 0 | T/T | 0 |
| HC772 | 0 | 0 | A/G | 1 | A/A | 0 | G/G | 0 | T/T | 0 |
| HC773 | 1 | 0 | G/G | 0 | A/A | 0 | T/G | 1 | C/T | 1 |
| HC774 | 0 | 0 | A/A | 2 | A/A | 0 | G/G | 0 | T/T | 0 |
| HC775 | 0 | 0 | G/G | 0 | G/A | 1 | G/G | 0 | T/T | 0 |
| HC776 | 0 | 0 | A/G | 1 | A/A | 0 | G/G | 0 | T/T | 0 |
| HC777 | 0 | 0 | A/G | 1 | G/A | 1 | G/G | 0 | T/T | 0 |
| HC778 | 0 | 0 | A/G | 1 | A/A | 0 | G/G | 0 | T/T | 0 |

| | | | | | | | | | | |
|---|---|---|---|---|---|---|---|---|---|---|
| HC779 | 0 | 0 | A/G | 1 | G/A | 1 | G/G | 0 | C/T | 1 |
